# The cost-effectiveness of testing and quarantine strategies to contain epidemic spread during the Hajj pilgrimage: A modelling study

**DOI:** 10.64898/2026.06.01.26354577

**Authors:** Jack Wardle, Anne Cori, Katharina Hauck, Pierre Nouvellet, Sangeeta Bhatia

## Abstract

The Hajj is an annual pilgrimage made by millions of Muslims to Mecca in the Kingdom of Saudi Arabia (KSA). The large number of international attendees at the Hajj increases the risk of global infectious disease spread. However, we know very little about the benefits, costs, and cost-effectiveness of testing and quarantining strategies to contain epidemic spread during mass gathering events.

In this work we developed a stochastic discrete-time compartmental metapopulation model to simulate international epidemics of infectious pathogens and their potential importation into KSA during the Hajj. We used the model and an epidemic simulation study to evaluate the impact and cost-effectiveness of three testing and quarantining strategies for arriving pilgrims: randomly testing 99% of pilgrims, 80% of pilgrims, or using a symptom-based screening strategy. The simulations lasted 100 days, covering the 30 days before the Hajj and 65 days after the Hajj.

Under the conditions assumed in our simulation study, there was strong evidence that testing and quarantining strategies are cost-effective measures for controlling epidemic threats at the Hajj. The median net monetary benefits of intervention strategies ranged from Intl$-41.89M [95% quantile range Intl$-42.37M–Intl$3.18B] to Intl$12.68B [Intl$-8.70B–Intl$13.82B] across scenarios with different pathogen characteristics (based on the natural histories of SARS-CoV-2 and H1N1 Influenza) and epidemic seed locations. Our results were sensitive to the data sources that were used to estimate the number of pilgrims travelling to KSA by origin country, with flight passenger statistics providing biased estimates of pilgrim numbers.

Our work provides an adaptable tool to inform infectious disease risk assessments and evaluate the cost-effectiveness of possible disease control measures for the Hajj, and could be extended to other mass gathering events.

## 1 Background

The Hajj is an annual pilgrimage made by Muslims to Mecca in the Kingdom of Saudi Arabia (KSA). The Hajj lasts for five days and is the largest annual mass gathering event in the world. For example, in 2019 approximately 2.4 million pilgrims attended the Hajj, comprised of 25% domestic and 75% foreign pilgrims [1, 2]. Although attendance reduced during and following the COVID-19 pandemic, pilgrim numbers are recovering (nearly 1.7 million attended in 2025) and the KSA government plans to increase the country’s capacity to accept pilgrims as part of their Vision 2030 program [3]. The Hajj contributes substantially to the KSA economy, with the accompanying revenues expected to exceed US$10 billion by 2030 [4]. Visas are required to attend and quotas are allocated to countries predominantly based on the size of their Muslim population, along with geopolitical factors [5].

As with any mass gathering event, the large international population movements associated with the Hajj increase the risk of global infectious disease spread. For instance, if the Hajj coincided with an epidemic or pandemic occurring elsewhere in the world, the arrival of large numbers of foreign pilgrims to KSA could lead to the introduction of a new pathogen (or further introductions of an already established pathogen) into KSA. This may lead to transmission between pilgrims and non-pilgrims, and risks of mortality and morbidity for both of these populations. There may be burdens on the KSA healthcare system, economic losses, and the need for potentially costly interventions.

Due to the potential risks of imported infectious diseases resulting in outbreaks, KSA authorities develop annual public health plans for the Hajj that identify infectious diseases of concern and disease-specific preventive measures to mitigate their impacts [6]. Interventions such as public health advice for high-risk groups, temperature checks for arriving pilgrims, or bans on travel from certain countries have been introduced for previous epidemics of severe acute respiratory syndrome (SARS), H1N1 influenza, Ebola, and SARS-Coronavirus-2 (SARS-CoV-2) [7–15].

Mathematical modelling can be used to help public health officials and decision-makers understand and manage infectious disease risks associated with pilgrim movements and the Hajj. For example, models were used to estimate the number of Middle East Respiratory Syndrome (MERS)-CoV cases that might occur among 2014 Hajj pilgrims under different incidence scenarios in KSA [16]. Previous modelling studies have also identified countries that may be at risk of importing H1N1 influenza or MERS-CoV following the return of pilgrims attending the Hajj in 2009 and 2012 respectively, and estimated the number of measles cases likely to be imported into KSA by Hajj pilgrims in 2019 [16–19].

Re-usable analytical and disease modelling frameworks could help decision-makers efficiently evaluate the risks of infectious disease outbreaks at each Hajj. Here we present a modelling study to help KSA decisionmakers evaluate the efficacy and cost-effectiveness of strategies for controlling epidemics imported during the Hajj. We developed a stochastic discrete-time compartmental metapopulation model that accounts for movement of pilgrims into and out of KSA, and simulated the importation of two respiratory pathogens by pilgrims arriving at the Hajj. We evaluated the epidemiological impact and cost-effectiveness of three testing protocols for arriving pilgrims, with the aim of helping decision makers select optimal testing strategies for mitigating infectious disease spread associated with the Hajj.

## 2 Methods

### 2.1 Epidemic model

#### 2.1.1 Model structure

Our stochastic discrete-time compartmental metapopulation model contains a set of patches that represent each home country from which Hajj pilgrims travel to KSA, and a set of patches representing Hajj pilgrims in KSA, split by their home country (where the home country could be KSA itself) (Figure S1). For simplicity, we assume that foreign pilgrims travel from their home countries to KSA and back, but there is no travel between other pairs of countries over the course of the simulation. We also include an additional patch to represent Saudi non-pilgrims that work at or live near the Hajj (referred to as ‘at risk’ non-pilgrims). We assume that the size of the ‘at risk’ patch is the sum of the populations of Mecca and Madinah.

Within each patch, we model the progression of individuals through different disease states (Figure 1). A new infection can occur upon contact between a susceptible (*S*) and an infectious person, with an assumption that the rate of mixing between individuals is 30% greater for pilgrims than non-pilgrims. After infection, individuals enter a latent period (*E*) and then become infectious. A proportion (*π*) of infected people develop symptoms, with the remainder being asymptomatic (*I*^*a*^). Individuals with symptomatic infections first pass through a state where they are presymptomatic but infectious (*I*^*p*^), followed by an infectious and symptomatic state (*I*^*s*^). It is assumed that presymptomatic and asymptomatic individuals have a lower infectiousness than symptomatic individuals. Infectious individuals then recover (*R*). Model equations and other assumptions are detailed in Suppl Sec. 1.1. Estimates of hospitalisations and deaths were computed in post-processing rather than modelled explicitly (see Section 2.2.1).

**Figure 1:**
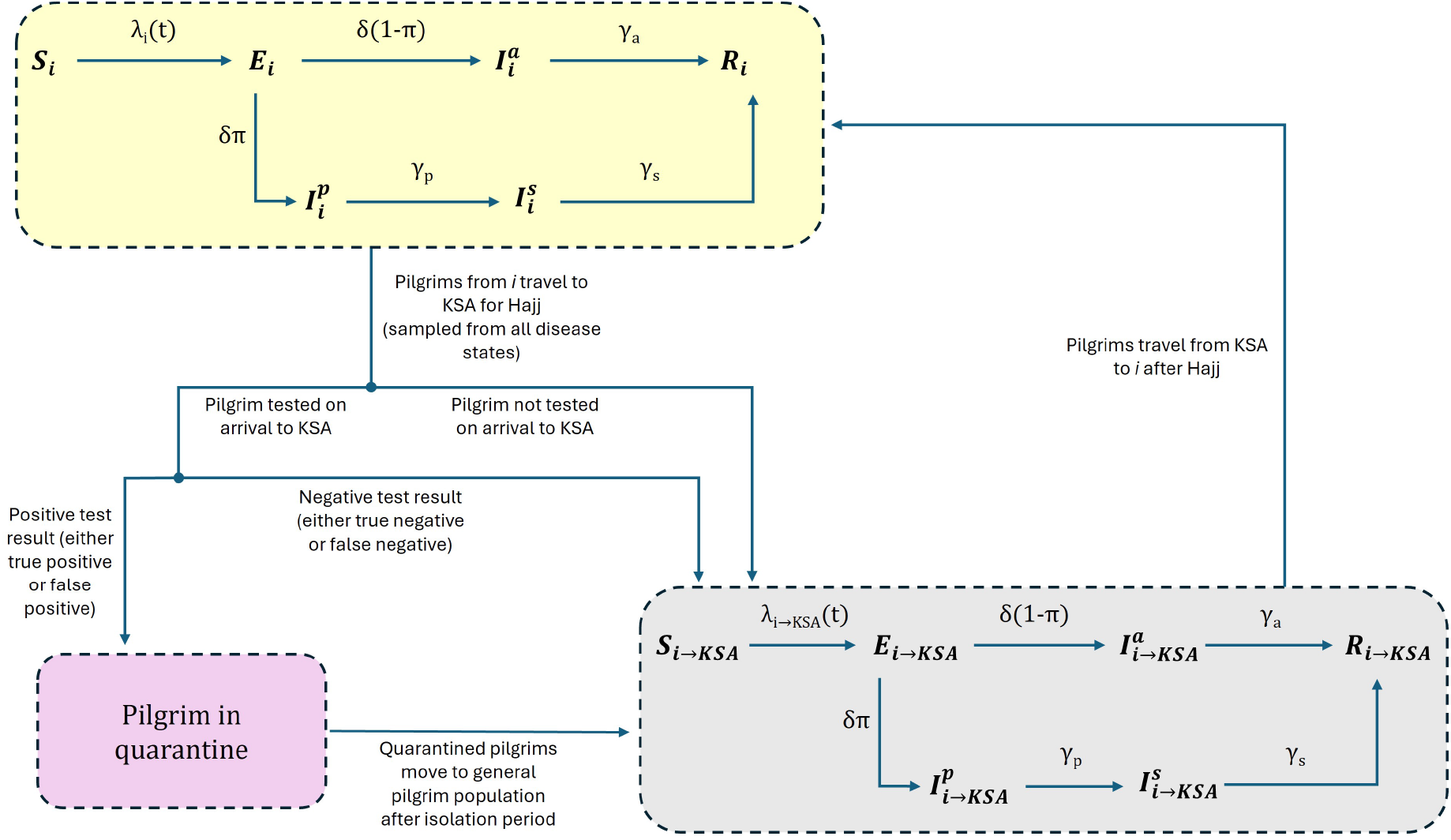
Transitions between compartments in model. The yellow rounded rectangle represents the disease transmission dynamics in the home countries (*i*) of pilgrims. Some of those individuals then travel to KSA to attend the Hajj during a defined 30-day period at the start of the simulation (non-pilgrims remain in the yellow rectangle for the duration of the simulation, only contributing to disease dynamics within their own country). If individuals are not tested on arrival, or receive negative test results, they immediately transition to the equivalent disease compartment in the grey rectangle, which denotes the disease transmission dynamics for the pilgrim population from country *i* in KSA (information on the origin of pilgrims is retained so that they travel back to their home countries at the end of the simulation). Individuals who receive a positive test result upon arrival must first transition through a quarantine stage (pink rectangle, in which information on *i* is retained) before moving in to the grey rectangle (in either the same or a later compartment to account for the duration of quarantine - see Suppl Sec. 1.1.3 for details). Following completion of the Hajj, pilgrims return to their original home countries. The subscript *i* → *KSA* denotes individuals from country *i* that are attending the Hajj in KSA. Other model symbols are defined in Eqs. (S1) to (S3). There are two populations that are treated differently: pilgrims from KSA proceed directly from the yellow to grey rectangle (bypassing any testing), while ‘at risk’ non-pilgrims are initially in the grey rectangle, before returning to the general KSA patch after the Hajj.

#### 2.1.2 Model steps

We ran our model simulations for 100 days (beginning 30 days before the Hajj started), which was sequentially composed of: 1) 30 days during which pilgrims arrived in KSA from their origin countries at a uniform rate; 2) 5 days during which the Hajj took place and there was no movement in or out of KSA; 3) 30 days during which pilgrims departed from KSA to their origin countries at a uniform rate; and 4) 35 day ‘post-Hajj’ period during which there was no movement in or out of KSA. We selected a 100-day simulation period under the assumption that effective vaccines or other pharmaceutical controls would be available after 100 days of an epidemic, in line with the targets of the Coalition for Epidemic Preparedness Innovations (CEPI) and other global health actors [20, 21].

At each time step (Δ*t* = 1 day) we modelled a sequence of three stochastic processes: 1) Pilgrim movement to/from KSA; 2) Testing/quarantining pilgrims arriving at the Hajj, and releasing any pilgrims who have finished quarantine; 3) Update of disease states (Figure 1). The mathematical details of each model step are provided in Suppl Sec. 1.1.3.

### 2.2 Simulation study

We simulated epidemics of respiratory pathogens resembling the characteristics of influenza and coronaviruses, and compared how epidemic outcomes varied under different strategies for the testing and quarantine of arriving pilgrims. We explored four testing/quarantining strategies in the simulations: i) No testing; ii) Intermediate testing (randomly testing 80% of arriving pilgrims); iii) High testing (randomly testing 99% of arriving pilgrims); iv) Symptom-based testing (also referred to as ‘syndromic’).

In the Intermediate and High testing strategies, we assumed use of a rapid antigen test with sensitivity of 0.7680 and a specificity of 0.9968, based on a SARS-CoV-2 test [22]. In the syndromic testing strategy, we assumed that pilgrims identified as having an elevated temperature would receive a further test. We assumed that this screening step would identify 87% of infected pilgrims and 39% of uninfected pilgrims for further testing, based on previous estimates of the effectiveness of airport thermal screening for influenza [23]. The pilgrims identified for further testing by thermal screening were then all tested with a rapid antigen test (same sensitivity and specificity as above). In our model, pilgrims receiving a positive result from the rapid antigen tests immediately entered quarantine facilities for a defined isolation period of 10 days.

We explored how the effectiveness of interventions might vary by pathogen type or by the location of the epidemic prior to its arrival in KSA (which can influence the number of imported infections). We varied the pathogen’s natural history parameters in the model to simulate epidemics for two pathogen scenarios: i) a pathogen with similar characteristics to H1N1 Influenza that we label as ‘Influenza-X’; ii) a pathogen with similar characteristics to SARS-CoV-2 that we label as ‘SARS-CoV-X’. The key parameters varied were basic reproduction number (Influenza-X: 1.64, SARS-CoV-X: 2.66), proportion of people that develop symptoms (Influenza-X: 0.669, SARS-CoV-X: 0.595), mean durations in days of different stages of infection (Influenza X: *E*=1.1, *I*^*p*^=1.0, *I*^*s*^=1.5, *I*^*a*^=2.5, SARS-CoV-X: *E*=4.6, *I*^*p*^=1.7, *I*^*s*^=2.1, *I*^*a*^=2.1). The full set of parameter values used in each pathogen scenario are shown in Suppl Tab. S2.

We simulated three epidemic location scenarios, where the epidemics coincided with the time of the Hajj: i) an emerging epidemic that was localised in Indonesia – the largest source of foreign pilgrims; ii) an emerging epidemic that was localised in the United Kingdom (UK) – selected as an example country that is a source of a relatively small number of foreign pilgrims; iii) epidemics that were established in multiple countries and continents (we refer to this as the Global scenario). In the Indonesia and UK scenarios, we began the simulations with 1,000 individuals infected. In the Global scenario, we seeded infections in the three most populated countries within each of the six World Health Organization regions (Suppl Sec. 1.5) and began the simulations with 0.001% of individuals infected in each of the 18 selected countries (corresponding to a total of almost 53,000 infections distributed globally at the beginning of the simulations).

Our primary analysis used historical pilgrim data from 2008 from the KSA Ministry of Health (MOH), reported by Khan *et al*, as a measure of the numbers and origins of pilgrims attending the Hajj [17]. More recent pilgrim data were not openly available for our use. We evaluated the sensitivity of our results to the use of mobility proxies (i.e. alternate data sources containing information on movement patterns) instead of MOH pilgrim estimates. We did this by running simulations where the numbers and origins of pilgrims were estimated using a flight passenger dataset from the International Air Transport Association (IATA) that contained the numbers of passengers that travelled between pairs of international airports each month in the period January 2012 to December 2021. We estimated pilgrim numbers from the IATA data either with or without adjusting for the baseline passenger levels that would be expected in the absence of the Hajj [24]. In the proxy with no baseline adjustment, we assumed that the proportions of pilgrims travelling from each country mirrored the proportions seen in the flight passenger data for August 2018 (the Hajj month in our data that had the most passenger arrivals in KSA). In the baseline-adjusted proxy, we subtracted the estimated baseline traffic (using arrival numbers observed in January 2012, the month with fewest arrivals in KSA in the IATA dataset) and assumed that the pilgrim proportions follow the distribution of origin countries among the remaining passengers. To ensure that pilgrim population sizes were comparable between movement scenarios, we scaled the pilgrim numbers estimated in the IATA methods so that the total pilgrim numbers were the same as reported in the MOH data. Additional details on the methods used to generate the IATA mobility proxies are outlined in Suppl Sec. 1.2.1.

All of the factors influencing epidemic outcomes that we explored are summarised in Table 1. The combination of scenarios across these four factors gave us a total of 72 epidemic scenarios. For each of these we simulated 1,000 epidemics over a 100-day period.

**Table 1:** Factors influencing epidemics that were explored in the simulation study. The scenarios that were considered under each factor are shown. See Suppl Sec. 1.2 for more details.

| Factor | Scenarios |
| --- | --- |
| Testing strategy | No testing; Intermediate testing/quarantine (*80% of pilgrims tested*); High testing/quarantine (*99% of pilgrims tested*); Symptom-based testing/quarantine |
| Pathogen | Influenza-X; SARS-CoV-X |
| Seed location | Global; Indonesia; United Kingdom |
| Movement input | MOH; IATA with adjustment for baseline passenger levels in absence of Hajj; IATA with no baseline adjustment |

#### 2.2.1 Summary metrics for simulations

For each epidemic simulation we computed: (1) The total number of cases in different sub-populations (i.e. foreign pilgrims, domestic pilgrims, domestic non-pilgrims). (2) The number of hospitalisations and deaths in sub-populations, by combining assumed age-sex distributions for the modelled populations with age-specific infection hospitalisation ratios (IHR) and infection fatality ratios (IFR) (Table S4). IHR values ranged from 1.60 × 10^−5^ – 1.84 × 10^−1^ and 1.83 × 10^−3^ – 1.03 × 10^−2^ for SARS-CoV-X and Influenza X respectively, while IFR values ranged from 1.60 × 10^−5^ – 7.80 × 10^−2^ and 7.36 × 10^−5^ – 6.56 × 10^−3^. We assumed that the ages and sex of all foreign pilgrims followed distributions reported by Azarpazhooh *et al* for a large cohort of Iranian pilgrims, with domestic pilgrims and non-pilgrims following estimates from United Nations World Population Prospects [25, 26]. (3) The years of life lost (YLL), by comparing the ages of those dying with the remaining life expectancy for a person of that age and sex. (4) The net monetary benefits (NMB) of testing strategies, which compare the costs and benefits of an intervention assuming a willingness to pay (WTP) threshold for a unit of health benefit [27]. In the main results we used a WTP threshold for KSA of Intl$20,324 for a YLL averted, based on values reported by Pichon-Riviere *et al* [28]. For NMB estimates we considered the YLLs averted among KSA residents (pilgrims and non-pilgrims), and the costs that arose from pilgrim testing/quarantine and hospital treatment of foreign pilgrims while in KSA and KSA residents. Cost estimates were based on data found in published literature and are summarised in Table S3. Further details of the summary metrics are provided in Suppl Sec. 1.2.5. In the main text we focus on presenting the YLL and NMB estimates, with estimates of cases, hospitalisations, and deaths shown in the Supplementary Material.

For each scenario, we summarised the distributions of each outcome across all 1,000 simulations using the median, 2.5%, 25%, 75%, and 97.5% quantiles. We paired simula-tions between the intervention (i.e. with testing) and baseline (i.e. no testing) scenarios to compute differences in YLLs and costs. The results presented in the main text are based on a ranked pairing method, whereby for each intervention scenario we first ordered the 1,000 simulations by the total number of cases occurring in KSA. We then paired each of the ranked simulations with the equal ranking simulation from the corresponding baseline scenario (matched on movement input, pathogen, and seed location), allowing us to compare epidemic trajectories to suitable counterfactuals, and limit misleading conclusions on the effects of interventions that can arise from pairing based on matching by initial random seeds [29]. In the Supplementary Material we include results from using a pairing method where the simulations were matched based on the value of an initial random seed within the simulation code.

### 2.3 Software

All epidemic simulations and analyses were performed using the R software [30]. The model code is available in the *multipatchr* package in R (https://github.com/sangeetabhatia03/multipatchr). The code for the analysis in this paper is available at https://github.com/j-wardle/hajj_modelling_study.

## 3 Results

Across combinations of pathogen and epidemic seed location in the central analysis de-scribed in the main text (where pilgrim flows were based on MOH pilgrim data), the median YLLs averted were largest for the High testing strategy, followed by the Syndromic and Intermediate testing strategies (Figure 2). However there was some overlap of 95% quantiles between testing strategies for all pathogen-location scenarios.

**Figure 2:**
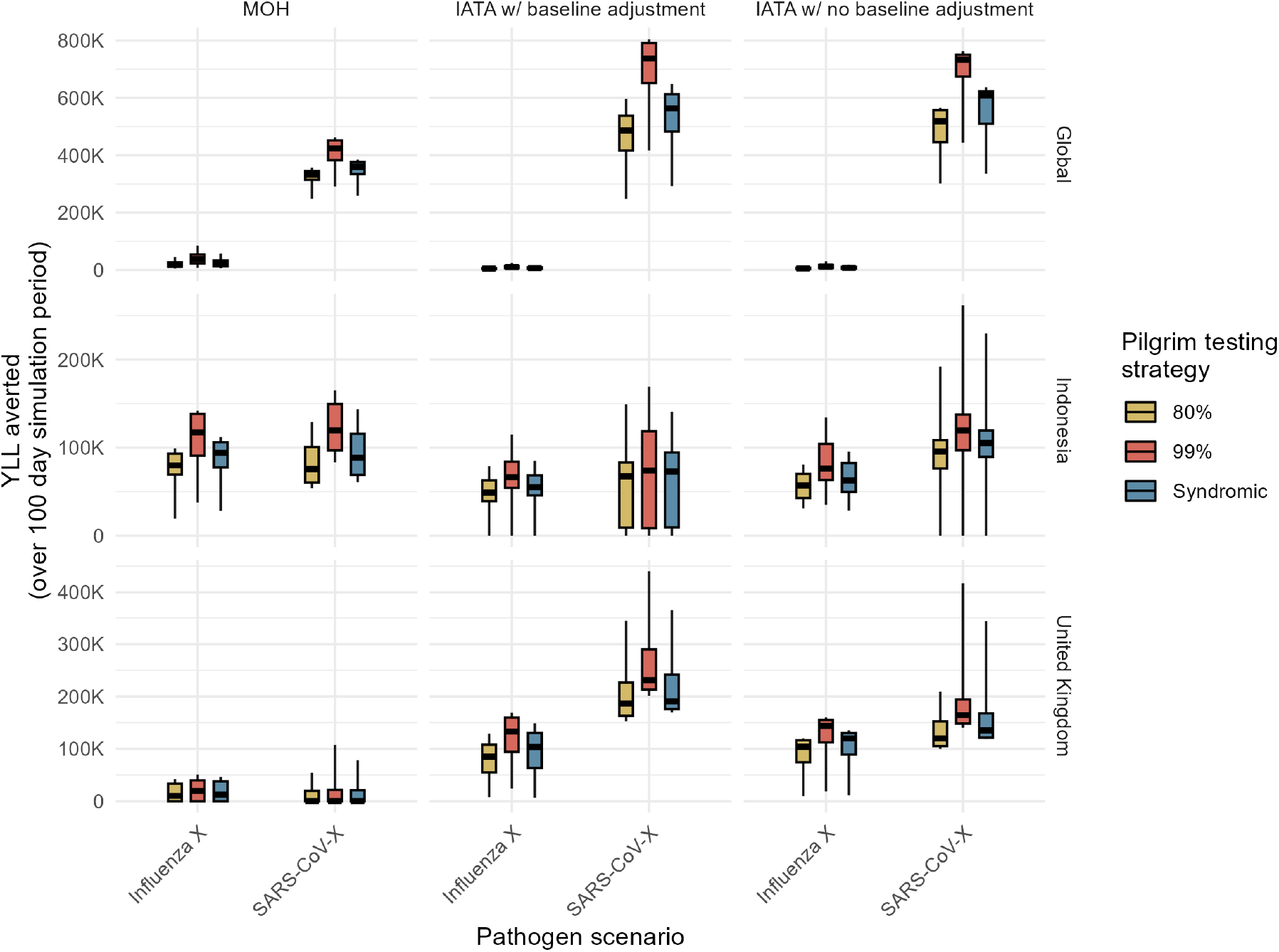
Years of life lost averted in KSA resident population through testing interventions. Years of life lost (YLL) averted are presented by movement input method (columns), epidemic seed scenarios (rows), and pathogen (x-axis). All values for YLL averted are estimated relative to the YLL in the equivalent scenarios with no testing of arriving pilgrims. Colours denote the testing strategy. Box and whisker plots summarise the median (thick black horizontal line), interquartile range (coloured rectangle) and 95% quantiles (range denoted by black lines) across 1,000 simulations.

The absolute differences in median YLLs averted between testing strategies, as well as the absolute values of the YLL averted, were smaller for the Influenza-X and SARS-CoV-X epidemics seeded in the UK. This reflected the small number of pathogen importation events among Hajj pilgrims in these scenarios (Figures S2 to S4). Despite differences in the numbers of imported infections and the overall effectiveness of testing strategies in averting YLLs, the numbers of people entering quarantine (and therefore the burden placed on quarantine facilities) were relatively similar across pathogen and epidemic location scenarios (Figures S2 to S4).

The interventions in our study were generally found to be cost-effective, with positive median NMBs for most testing strategies across pathogen and epidemic seed scenarios (Figure 3). A positive NMB means that the benefits of the intervention are greater than their costs at the assumed WTP threshold. Testing/quarantining interventions were most cost-effective for SARS-CoV-X scenarios where there was already some widespread global transmission prior to the Hajj. In contrast, the only testing strategies with negative median NMB estimates were the UK SARS-CoV-X scenarios, although the values were relatively small and the uncertainty estimates crossed zero indicating that the intervention was cost-effective in some simulations. The negative NMB estimates for the UK SARS-CoV-X scenarios using the MOH data were driven by the fact that, even in the absence of interventions, many of the simulations in the ‘no testing’ scenario did not lead to epidemics within KSA because there were no imported infections (Figure S3). Consequently, the introduction of testing interventions had no effect in averting lives lost, because there were no years of life lost in either the ‘no testing’ or intervention scenario. Generally, the differences between testing strategies within pathogen-seed location combinations were smaller than the differences that occurred when pathogen or seed location were varied.

**Figure 3:**
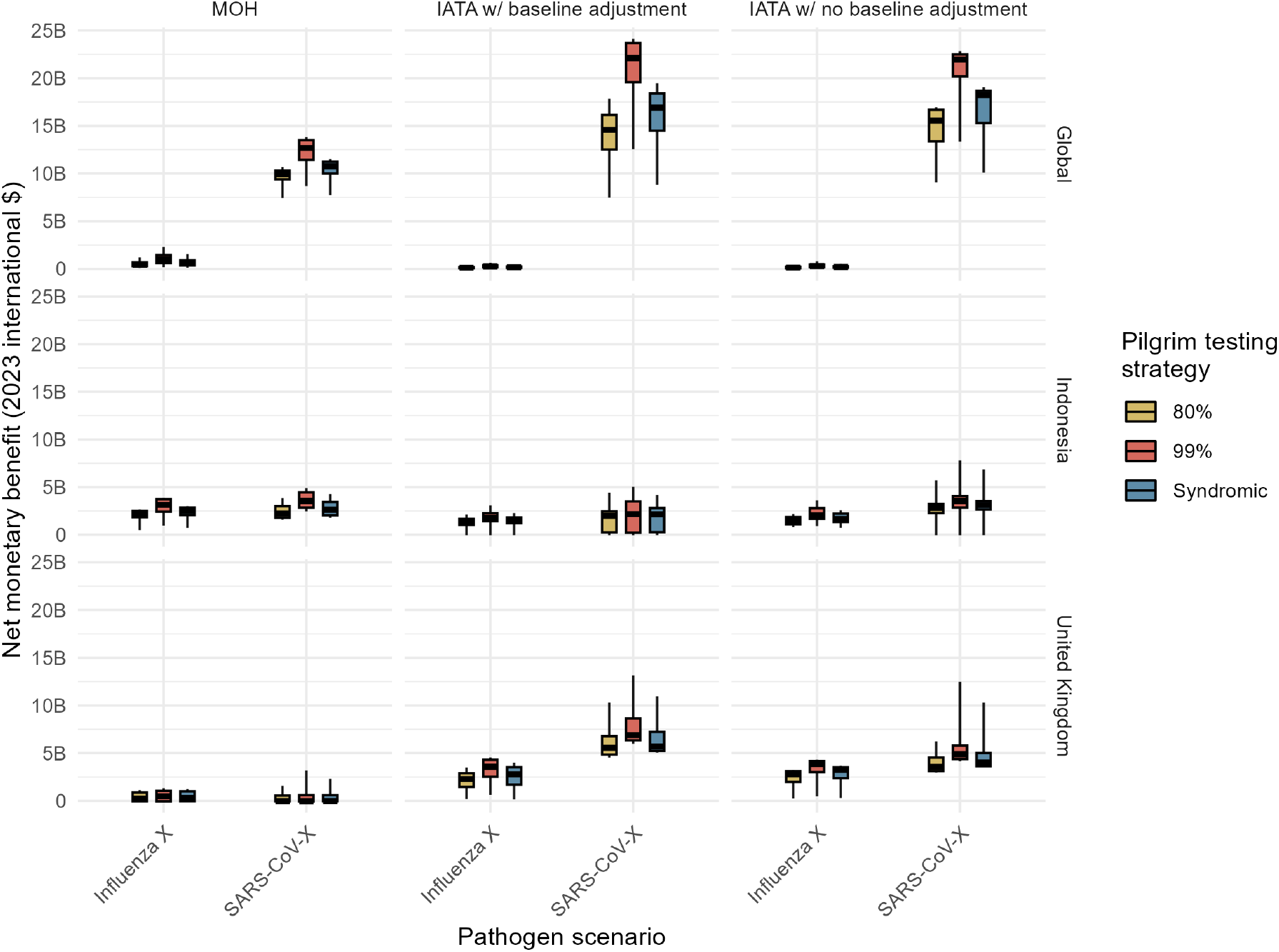
Net monetary benefit estimates for different intervention scenarios, relative to a baseline scenario with no testing. Estimates of the net monetary benefit (NMB) are provided in 2023 international $. The approach used to estimate NMB values is described in Suppl Sec. 1.2.5. NMB estimates are presented by movement input method (columns), epidemic seed scenarios (rows), and pathogen (x-axis). Colours denote the testing strategy. Box and whisker plots summarise the median (thick black horizontal line), interquartile range (coloured rectangle) and 95% quantiles (range denoted by black lines) across 1,000 simulations.

There were substantial differences between pathogen-epidemic seed scenarios in the NMB estimates for each testing strategy, with the largest difference between the High testing strategy in the Global SARS-CoV-X scenario and the equivalent testing strategy in the UK SARS-CoV-X scenario (median NMBs of Intl$12.68B and Intl$-41.89M respectively). However, within-scenario differences between the median NMBs of testing strategies were relatively small (the maximum within-scenario difference was Intl$2.7B for the Global SARS-CoV-X scenario). The probabilities that interventions were cost-effective were generally unaffected by changes in the WTP threshold (Figure S16).

Our results showed some sensitivity to the use of mobility proxies. For example, NMBs of the testing/quarantining strategies in the UK SARS-CoV-X scenarios were evaluated as being large and positive with both IATA mobility proxies, whereas estimates were negative with the MOH pilgrim data. The differences stem from the considerable variation in the numbers and origins of pilgrims between the different mobility proxies (Figure 4; the total number of pilgrims was the same across all the mobility proxies we compared due to scaling of the IATA data). The number of pilgrims attending the Hajj as reported by the MOH were substantially different from those estimated using IATA data. For instance, while Indonesia was the largest source of pilgrims in the MOH data, it was not classified as a high-contribution country (defined as being part of the smallest subset of countries that together contribute ≥ 80% of foreign pilgrims) for IATA data (irrespective of the adjustment method used). In fact, using the IATA data resulted in >80% fewer pilgrims from Indonesia compared with the MOH data. Similarly, the numbers of pilgrims attending from Iran, Nigeria, and Algeria were vastly underestimated by the IATA data compared to MOH data. In contrast, IATA data overestimated the numbers of pilgrims travelling from some countries. The number of pilgrims expected to arrive from Egypt was increased by 240% and 310% for the two IATA methods. The United Arab Emirates and UK were expected to be sources of a considerable number of pilgrims based on the IATA data, but comparison with the MOH numbers suggested that this was a large overestimation. Applying the simple baseline adjustment approach to the IATA data did not improve estimates of pilgrim numbers, and in fact generally gave greater discrepancies versus MOH estimates than the IATA input with no adjustment.

**Figure 4:**
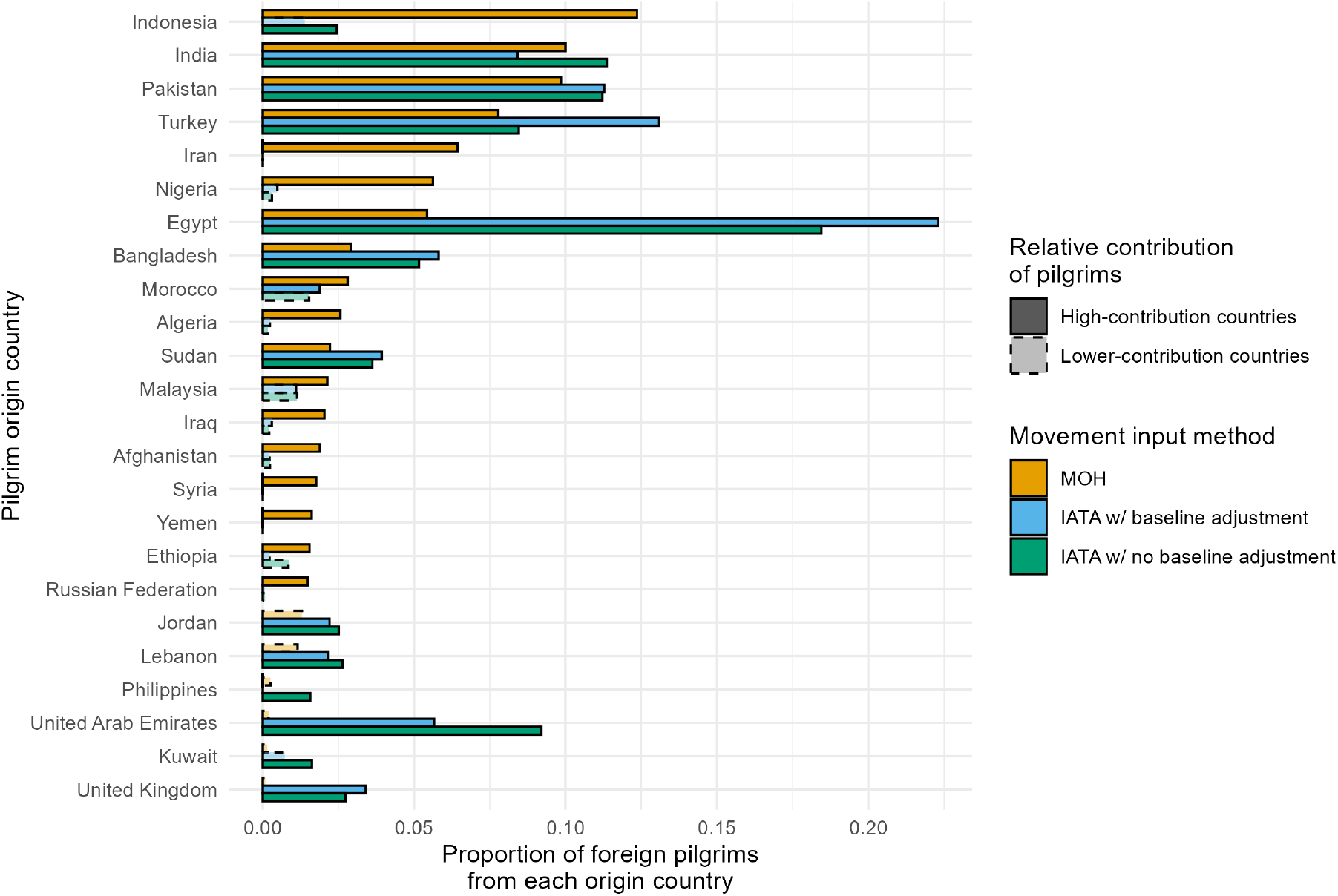
Comparison of pilgrim movement input methods. The bars show the proportion of foreign pilgrims travelling from each origin country. A high-contribution country (bars with darker shading/solid outlines) is one of the countries that send the most pilgrims for a given movement input method, defined as being part of the smallest subset of countries that together contribute ≥ 80% of foreign pilgrims; remaining countries are classed as lower-contribution countries (lighter shading/dashed outlines). We present in the figure the set of countries that are high-contributors for at least one of the movement input methods. Colours represent the three movement input methods; MOH is Kingdom of Saudi Arabia Ministry of Health data, and IATA are flight passenger data (see Suppl Sec. 1.2.1 for details of the baseline adjustment methods).

The results presented here are all based on a simulation period of 100 days. To assess the sensitivity of our findings to the length of the simulation, we ran some additional simulations with a 300-day simulation period. We found that the longer we run simulations for following the initial importation period, the more similar the different scenarios were (Figure S20).

## 4 Discussion

We designed an infectious disease transmission model that was parameterised with esti-mates of pilgrim movements to the Hajj and used to evaluate the potential impact and cost-effectiveness of testing and quarantine strategies for mitigating the risks of imported epidemics. Under the conditions assumed in our simulation study, there was strong evidence that testing and quarantining strategies are cost-effective measures for controlling epidemic threats at the Hajj. Interventions were most cost-effective for the SARS-CoV-X scenario where there was already transmission occurring in multiple countries prior to the Hajj. This was because i) more deaths were averted in the SARS-CoV-X scenarios (SARS-CoV-X had greater IFRs than Influenza X); and ii) testing/quarantine in the global scenarios prevented more importations of infections (compared to scenarios with smaller initial epidemics in Indonesia and United Kingdom).

The high NMBs in our analysis are likely being driven by the high costs of hospital treatment that are avoided through interventions, relative to the costs of implementing screening, as well as the deaths that are averted. The respective costs of testing and treatment are therefore key variables (alongside the WTP threshold), and highlight the importance of accurate costing data for informing decision-making. We also assumed that the costs of running an airport screening programme increased linearly with the number of passengers. A non-linear cost function (with high set-up costs, and then lower marginal costs for each additional passenger screened) may be more realistic. This is for future research, subject to the availability of better costing data.

The epidemic dynamics over the time period we looked at are fundamentally linked to the number of importations that occur. Scenarios with more importations (either through increased arrivals from the seed locations, or greater prevalence of infection among arriving pilgrims) resulted in greater epidemic sizes. The effect of interventions in the model is dependent on the length of time that we run the simulations for, and the choice of time period may therefore depend on the exact priorities of policymakers. For example, if officials’ main interests are in the extra burden of cases, hospitalisations and deaths in KSA from foreign pilgrims, it may be more appropriate to run simulations for a shorter time period of 65 days, since this would cover the approximate duration of pilgrim stays (although it could be argued that hospitalisation may prolong someone’s stay). A similar time period may also be appropriate if the interest was in the number of exportations that could occur from foreign pilgrims leaving KSA after the Hajj.

The ‘100 days mission’ is an ambitious and worthwhile target in global health. Advancements in vaccine technology and manufacturing capacity mean that we are in a better place to achieve this target than before the SARS-CoV-2 pandemic, but there are no guarantees that future epidemics would reach a controllable stage after 100 days, as has been seen with the ongoing mpox epidemic [31]. We ran some additional simulations with a 300-day simulation period. Conditional on some disease importations occurring when pilgrims arrive in KSA, in the long-term the epidemic outcomes were similar between different testing scenarios, including the no testing scenario. This emphasises that interventions that delay or slow the importation of new pathogens are likely to only be worthwhile if the additional time is used to introduce effective control measures, in line with the findings of previous research [32–36].

Some of our findings contrast with other modelling studies that have suggested that arrival screening is likely to miss the majority of infectious cases [37, 38]. This can be partly explained by our assumption that the sensitivity of the diagnostic test is the same across all infected states. If we instead assumed that the test was less sensitive for exposed, asymptomatic, or presymptomatic pilgrims, we would expect more false negative test results and therefore the interventions would be less effective. A full exploration of the assumptions around test sensitivity and intervention costs was outside the scope of this current work. However, our work has established a modelling framework, with publicly available code, that can be modified to allow assessment of these factors in the future, ideally in partnership with local decision-makers.

The importance of collaboration with local decision-makers is further emphasised by our results that highlighted some of the data gaps limiting the use of infectious disease models for health planning at mass gathering events. Detailed statistics providing the numbers and origins of pilgrims that travel to KSA for the Hajj are typically unavailable to the research community. We found that, in the absence of official MOH data, the use of mobility proxies can have substantial impacts on modelled epidemic outcomes and the estimated cost-effectiveness of disease control interventions. Although flight passenger data are a common data source in models of international disease spread [32, 39–43], in this work we showed that these data can be misleading for modelling the introduction of epidemic diseases into KSA during the Hajj. Since many pilgrims travel to KSA on charter flights, the IATA data (which does not include charter flights) gives biased estimates of number of pilgrims travelling to KSA for the Hajj from each origin country. Official statistics of pilgrim numbers by origin are occasionally shared with researchers (e.g. [17]). Our work emphasises the importance of modellers having access to these data in order to accurately evaluate the risks posed by epidemic pathogens that may be imported by foreign pilgrims travelling to the Hajj, and to support KSA decision-makers in finding optimal interventions to reduce morbidity and mortality for pilgrims from across the world.

We have demonstrated how our epidemic model for mass gatherings could be applied to evaluate the benefits and costs of public health interventions to reduce epidemic risks at mass gathering events, and we share the methodology underpinning the model framework as well as the underlying model code. We note that though the results presented here are for a specific set of scenarios and assumptions, they can provide some insights into the potential effects of parameters such as IHR, IFR, and importation rates on cost-effectiveness estimates. However, we would advise caution in generalising our findings to other scenarios without first running further simulations to test the sensitivity of our results to some of the assumptions made. The assumptions that merit further testing include: i) the level of mixing in pilgrim populations relative to non-pilgrims; ii) who falls in the ‘at risk’ population; iii) accuracy of cost estimates (limited data were available on the costs of operating an arrival testing programme, and none from a KSA setting); iv) age-sex distributions of pilgrims (changes in demographics will influence hospitalisation estimates due to the age-specific IHR we used); v) varying diagnostic sensitivity by infection state.

We have presented simulation scenarios here to show how our model could be used, and future work could focus on developing the model into a user-friendly R package or other interface to be used by policymakers working on the Hajj or other mass gathering events. In any future development, the model should ideally be validated against disease surveillance data from the Hajj. Our model can also be easily adapted to explore additional testing strategies (e.g. only testing pilgrims arriving from countries with local transmission) and other pathogen scenarios (e.g. for a pathogen with a larger proportion of asymptomatic transmission). As such, our work provides a useful foundation for assisting public health planning for the Hajj, as well as other international mass gathering events, and supporting the mitigation of epidemic threats.

## Data Availability

The model code is available in the multipatchr package in R (https://github.com/sangeetabhatia03/multipatchr). The code for the analysis in this paper is available at https://github.com/j-wardle/hajj_modelling_study.

## Acknowledgements

JW acknowledges research funding from the Wellcome Trust (grant 102169/Z/13/Z). AC acknowledges the Academy of Medical Sciences Springboard, funded by the Academy of Medical Sciences, Wellcome Trust, the Department for Business, Energy and Industrial Strategy, the British Heart Foundation, and Diabetes UK (reference SBF005*\*1044). KH acknowledges funding by Kenneth C Griffin. SB acknowledges funding by the Wellcome Trust (223120/Z/21/Z). AC and KH acknowledge funding from the National Institute for Health Research (NIHR) Health Protection Research Unit in Health Analytics & Modelling (NIHR207404), a partnership between UK Health Security Agency (UKHSA), Lon-don School of Hygiene & Tropical Medicine, and Imperial College of Science, Technology, & Medicine. JW, AC, KH and SB acknowledge funding from the MRC Centre for Global Infectious Disease Analysis (reference MR/X020258/1), funded by the UK Medical Re-search Council (MRC). This UK funded award is carried out in the frame of the Global Health EDCTP3 Joint Undertaking. JW and KH acknowledge funding by Community Jameel. Disclaimer: The views expressed are those of the author(s) and not necessarily those of the NIHR, UK Health Security Agency or the Department of Health and Social Care.

