## Supplementary material for "The cost-effectiveness of testing and quarantine strategies to contain epidemic spread during the Hajj pilgrimage: A modelling study"

**2 Supplementary results** **15**

### 1 Supplementary methods

#### 1.1 Epidemic model

##### 1.1.1 Spatial structure

The model contains a set of patches that represent each home country from which Hajj pilgrims travel to KSA, and a set of patches representing Hajj pilgrims in KSA, split by their home country (where the home country could be KSA itself) (Fig. S1). For simplicity, we assume that foreign pilgrims travel from their home countries to KSA and back, but there is no travel between other pairs of countries over the course of the simulation. We also include an additional patch to represent Saudi non-pilgrims that work at or live near the Hajj (we refer to this population as 'at

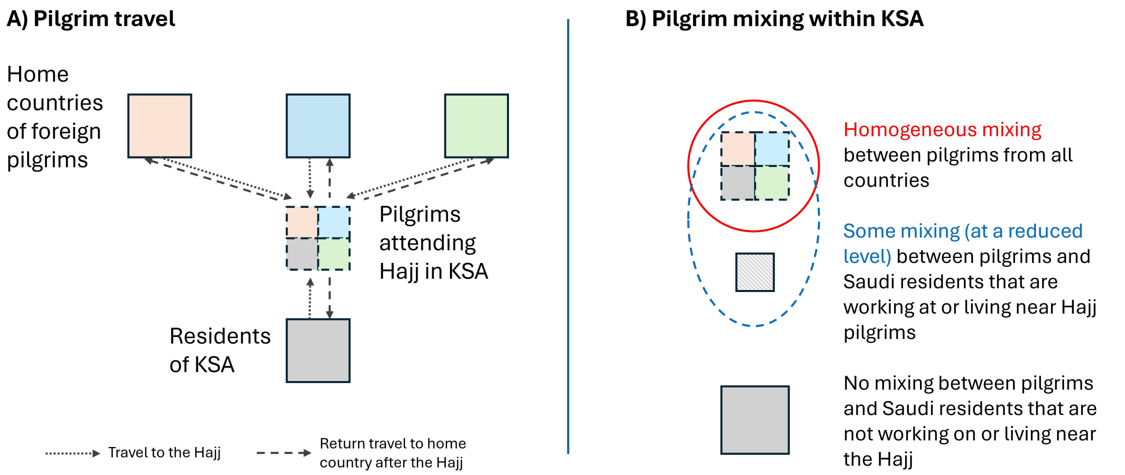

Figure S1: **Pilgrim movement and mixing patterns in the metapopulation model.** A) Large coloured squares represent the populations in the home countries of Hajj pilgrims, with grey representing the population in Kingdom of Saudi Arabia (KSA) and other colours representing the home countries of foreign pilgrims. Smaller squares indicate the pilgrim populations from each of these countries, that travel to and from KSA. B) The assumptions of mixing between different populations within KSA.

risk' non-pilgrims). We assume that the size of the 'at risk' patch was the sum of the populations of Mecca and Madinah (the primary locations visited by Hajj pilgrims).

##### 1.1.2 Progression of infection

Within each patch, we model the progression of individuals through different disease states (susceptible, exposed, asymptomatic infectious, presymptomatic infectious, symptomatic infectious, recovered) (Figure 1 - main text). New infections occur upon contact between a susceptible ($S$) and infectious person. After infection, individuals enter a latent period ($E$) and then become infectious. A proportion ($\pi$) of infected people develop symptoms, with the remainder being asymptomatic ($I^a$). Individuals with symptomatic infections first pass through a state where they are presymptomatic but infectious ($I^p$), followed by an infectious and symptomatic state ($I^s$). Infectious individuals then recover ($R$). The transitions in the time interval $t + \Delta t$ are defined by the following equations:

$$[S_x \rightarrow E_x](t) \sim Binomial\left(S_x(t), 1 - \exp\left(-\lambda_x(t)\Delta t\right)\right) \tag{S1}$$

$$\begin{aligned}
&[E_x \rightarrow (I_x^a \ or \ I_x^p)](t) = N_x^{inf}(t) \sim Binomial\left(E_x(t), 1 - \exp\left(-\delta\Delta t\right)\right)\\
&Number \ of \ I^p \ in \ N_x^{inf}(t) = X(t) \sim Binomial\left(N_x^{inf}(t), \pi\right)\\
&Number \ of \ I^a \ in \ N_x^{inf}(t) = N_x^{inf}(t) - X(t)\\
&\therefore [E_x \rightarrow I_x^p](t) = X(t)\\
&and \ [E_x \rightarrow I_x^a](t) = N_x^{inf}(t) - X(t)
\end{aligned} \tag{S2}$$

$$\begin{aligned}
[I_x^p \to I_x^s](t) &\sim Binomial\left(I_x^p(t), 1 - \exp\left(-\gamma_p \Delta t\right)\right) \\
[I_x^a \to R_x](t) &\sim Binomial\left(I_x^a(t), 1 - \exp\left(-\gamma_a \Delta t\right)\right) \\
[I_x^s \to R_x](t) &\sim Binomial\left(I_x^s(t), 1 - \exp\left(-\gamma_s \Delta t\right)\right)
\end{aligned} \tag{S3}$$

where $1/\delta$ is the mean duration of the latent period, and $1/\gamma_a$, $1/\gamma_p$, and $1/\gamma_s$ are the mean durations of the asymptomatic, presymptomatic, and symptomatic infectious periods respectively. $x$ denotes the patch being modelled.

In the model, we assume that the extent of mixing, and therefore the force of infection ($\lambda_x$), depends on the population sub-group that the patch $x$ belongs to. These sub-groups are: i) pilgrims, both domestic and foreign, attending the Hajj (for whom the force of infection is represented by $\lambda_{pilgrim}(t)$); ii) non-pilgrims, both domestic and foreign, not attending the Hajj (force of infection in each country $i$ represented by $\lambda_{non-pilgrim,i}(t)$); and iii) 'at risk' non-pilgrims (force of infection represented by $\lambda_{AtRisk}(t)$). The forces of infection acting on each of the three sub-groups at time $t$ are computed as below:

$$\begin{aligned}
\lambda_{pilgrim}(t) &= \frac{\beta_{pilgrim} \sum_i \left(\theta_a I^a_{i\to KSA}(t) + \theta_p I^p_{i\to KSA}(t) + I^s_{i\to KSA}(t)\right)}{N_{AtRisk}(t) + \sum_i N_{i\to KSA}(t)} \\
&\quad + \frac{\beta \left(\theta_a I^a_{AtRisk}(t) + \theta_p I^p_{AtRisk}(t) + I^s_{AtRisk}(t)\right)}{N_{AtRisk}(t) + \sum_i N_{i\to KSA}(t)} \\
\lambda_{non-pilgrim,i}(t) &= \frac{\beta \left(\theta_a I^a_i(t) + \theta_p I^p_i(t) + I^s_i(t)\right)}{N_i(t)} \\
\lambda_{AtRisk}(t) &= \frac{\beta \left(\theta_a I^a_{AtRisk}(t) + \theta_p I^p_{AtRisk}(t) + I^s_{AtRisk}(t)\right)}{N_{AtRisk}(t) + \sum_i N_{i\to KSA}(t)} \\
&\quad + \frac{\beta \sum_i \left(\theta_a I^a_{i\to KSA}(t) + \theta_p I^p_{i\to KSA}(t) + I^s_{i\to KSA}(t)\right)}{N_{AtRisk}(t) + \sum_i N_{i\to KSA}(t)}
\end{aligned} \tag{S4}$$

where $\beta_{pilgrim}$ represents the per capita transmission rate between pilgrims, and $\beta$ is the per capita transmission rate for any other pair (i.e. where there is at least one non-pilgrim). $\theta_a$ is the infectiousness of an asymptomatic infectious case and $\theta_p$ is the infectiousness of a presymptomatic case, both relative to the infectivity of a symptomatic infectious case. The subscript $i \to KSA$ denotes individuals from country $i$ that are attending the Hajj in KSA, so that $I^a_{i\to KSA}(t)$, $I^p_{i\to KSA}(t)$, and $I^s_{i\to KSA}(t)$ represent the number of infectious individuals attending the Hajj from $i$ at time $t$ that are asymptomatic, presymptomatic, and symptomatic respectively. In a similar manner, subscript $i$ represents individuals in country $i$, while the subscript $AtRisk$ is used for individuals located in the 'at risk' population (i.e. $I^s_i(t)$ and $I^s_{AtRisk}(t)$ are the number of people who are symptomatic and infectious at time $t$ that are located in $i$ or $AtRisk$ respectively). $N_i$ is the number of people in country $i$, $N_{AtRisk}$ is the number of 'at risk' non-pilgrims, and $N_{i\to KSA}$ is the number of pilgrims from country $i$ in KSA. Since some individuals travel from $i$ to $KSA$ (and back) over time, the size of $N_i$ and $N_{i\to KSA}$ depends on $t$.

We used $\beta_{pilgrim}$ in the model to account for potentially higher infection rates between pilgrims due to high levels of crowding in pilgrim camps and at pilgrimage sites (the density of crowds at the Hajj can reach seven people per m$^2$ [1]). The value of $\beta_{pilgrim}$ was set by applying a multiplier to $\beta$. There were no studies that quantified the reproduction number of infectious diseases among Hajj pilgrims relative to general populations. Therefore, we quantified the potential effect of crowding on transmission rates using previous estimates of increases in reproduction numbers for diphtheria outbreaks in Bangladeshi refugee camps [2, 3]. The researchers found that $R_0$ was 1.65 times higher in refugee camps compared to other settings [3]. We assumed that the increase in respiratory disease transmission between pilgrims would take a value greater than the general population but lower than the refugee camp estimate. We therefore obtained values for $\beta_{pilgrim}$ by applying an assumed multiplier of 1.3 to $\beta$.

###### 1.1.3 Model steps

We ran our model simulations for 100 days (beginning 30 days before the Hajj started), which was sequentially comprised of:

1. 30 days during which pilgrims arrived in KSA from their origin countries at a uniform rate;
2. 5 days during which the Hajj took place and there was no movement in or out of KSA;
3. 30 days during which pilgrims departed from KSA to their origin countries at a uniform rate; and
4. 35 day 'post-Hajj' period during which there was no movement in or out of KSA.

At each time step ($\Delta t = 1$ day) we modelled a sequence of three stochastic processes: 1) Pilgrim movement to/from KSA; 2) Testing/quarantining pilgrims arriving at the Hajj, and releasing any pilgrims who have finished quarantine; 3) Update of disease states.

**<u>STEP 1: Pilgrim movement to/from KSA</u>**

For $t \leq 30$ days, we modelled the movement of Hajj pilgrims from their home countries to KSA. The daily number of pilgrims arriving in KSA from each disease compartment in country $i$ ($X^{travel}_{i \to KSA}$) was given by:

$$\begin{aligned}&\left[S_i \to S^{travel}_{i\to KSA}, E_i \to E^{travel}_{i\to KSA}, \ldots, R_i \to R^{travel}_{i\to KSA}\right](t)\\ &\sim Multinomial\left(pilgrims_i(t), prop\_S_i(t), prop\_E_i(t), \ldots, prop\_R_i(t)\right)\end{aligned} \tag{S5}$$

where $pilgrims_i(t)$ is the number of pilgrims travelling from country $i$ to KSA at time $t$ (see Section 1.2.1). $prop\_X_i(t)$ is the proportion of people in country $i$ that are in disease compartment $X$ at time $t$ (where $X = S, E, I^a, I^p, I^s, R$).

For $35 < t \leq 65$ days, we modelled the return of pilgrims to their home countries from KSA following the completion of the Hajj. The daily number of pilgrims returning from KSA to $i$ from each disease compartment was given by:

$$[S_{i\to KSA} \to S_i, E_{i\to KSA} \to E_i, \ldots, R_{i\to KSA} \to R_i]\,(t)$$
$$\sim Multinomial\,(pilgrims_i(t), prop\_S_{i\to KSA}(t), prop\_E_{i\to KSA}(t), \ldots, prop\_R_{i\to KSA}(t)) \tag{S6}$$

where $prop\_X_{i\to KSA}(t)$ is the proportion of pilgrims in KSA from country $i$ that are in disease compartment $X$ at time $t$. As noted above, there was no movement of individuals between patches for $30 < t \leq 35$ and $t > 65$ days.

#### STEP 2: Testing and quarantine

For $t \leq 30$ days, a proportion of the foreign pilgrims that arrive in KSA underwent testing. We modelled two approaches: random testing (detailed here), and a modified testing approach that focused on symptomatic travellers (see Section 1.2.4 below for details).

In the random testing strategy, arriving pilgrims ($X^{travel}_{i\to KSA}$) were randomly tested with a rapid diagnostic test at a defined testing rate (Eq. (S7)). Of those tested, pilgrims that received a true or false positive diagnosis (dependent on the defined sensitivity and specificity of the diagnostic test) immediately entered quarantine facilities for a defined fixed isolation period ($\phi$). The numbers entering quarantine were determined by the true positive and false positive diagnoses as outlined in Eqs. (S8) and (S9). All arriving pilgrims that were not tested or received a negative test result progressed to their current disease compartment in the patch for pilgrims from the same home country, $X_{i\to KSA}$ (Figure 2).

***Tested on arrival***

$$X^{tested}_{i\to KSA}(t) \sim Binomial\left(X^{travel}_{i\to KSA}(t), \tau\right) \tag{S7}$$
$$\text{where } X = S, E, I^a, I^p, I^s, R$$

where $\tau$ is the proportion of arriving pilgrims that are tested, $X^{travel}_{i\to KSA}(t)$ is the number of pilgrims arriving in KSA from country $i$ on day $t$ that are in disease compartment $X$, and $X^{tested}_{i\to KSA}$ is the number of pilgrims arriving from $i$ that are tested on arrival.

***True positive diagnoses on arrival***

$$X^{diagnosed}_{i\to KSA}(t) \sim Binomial\left(X^{tested}_{i\to KSA}(t), \epsilon\right) \tag{S8}$$
$$\text{where } X = E, I^a, I^p, I^s$$

where $\epsilon$ represents the test sensitivity.

***False positive diagnoses on arrival***

$$X^{false\_diagnosed}_{i\to KSA}(t) \sim Binomial\left(X^{tested}_{i\to KSA}(t), \mu\right) \tag{S9}$$
$$\text{where } X = S, R$$

where $\mu$ represents the false positive rate, calculated as 1 - test specificity.

***Pilgrims leaving quarantine***

At each time step we computed the number of pilgrims that reached the end of their quarantine (assumed to start immediately after a positive test and to be of fixed duration $\phi$) and were released back to the general population. The disease states ($S$, $E$, $I^a$, $I^p$, $I^s$, $R$) of the pilgrims released at time $t$ depended on: i) the number of pilgrims in each disease state that entered quarantine isolation at time $t - \phi$; ii) the probabilities that an individual who entered quarantine in a given disease state was in any of the possible subsequent disease states at the end of their quarantine period (for example, the probability that an individual who entered quarantine in $E$ was in $E, I^a, I^p, I^s$ or $R$ at the end of their quarantine). We computed the probabilities using a numerical simulation approach. For each possible starting disease state, we ran 1,000,000 simulated random disease progression pathways for a duration $\phi$, and recorded the proportion of simulations where the final disease state was $S$, $E$, $I^a$, $I^p$, $I^s$, or $R$. Individuals who entered quarantine in $S$ or $R$ (following a false positive diagnosis) remained in those disease states for the duration of their quarantine. Therefore, the number of people released to $S_{i \to KSA}$ and $R_{i \to KSA}$ at time $t$ was given by:

$$[X_{i \to KSA}^{false\_diagnosed} \to X_{i \to KSA}](t) = X_{i \to KSA}^{false\_diagnosed}(t - \phi) \tag{S10}$$

where $X = S, R$

The numbers and disease states of pilgrims released from quarantine at time $t$ for all other starting disease states were given by:

$$\begin{aligned}[X_{i \to KSA}^{diagnosed} \to (E_{i \to KSA}, I_{i \to KSA}^{a}, I_{i \to KSA}^{p}, I_{i \to KSA}^{s}, R_{i \to KSA})](t) \\ \sim Multinomial(X_{i \to KSA}^{diagnosed}(t - \phi), \\ p\_E_X^{qend}, p\_I_X^{a\_qend}, p\_I_X^{p\_qend}, p\_I_X^{s\_qend}, p\_R_X^{qend})\end{aligned} \tag{S11}$$

where $X = E, I^a, I^p, I^s, R$

and where notation of the form $p\_E_X^{qend}$ represents the probability that an individual who began their quarantine in disease state $X$ would end their quarantine in state $E$ (and so on for the other probabilities $p\_I_X^{a\_qend}$, $p\_I_X^{p\_qend}$, $p\_I_X^{s\_qend}$, and $p\_R_X^{qend}$), as computed by the numerical simulation approach described above.

**STEP 3: Update disease states**

The model then updated the disease states in each patch using Eqs. (S1) to (S4).

#### 1.2 Simulation study

We conducted a simulation study to compare how epidemic outcomes among pilgrim and KSA residents differed across a range of scenarios. The scenarios explored how varying the following factors affected the predicted epidemic outcomes: 1) the mobility proxy used to estimate the number of foreign pilgrims travelling to KSA from each country; 2) the characteristics of the epidemic pathogen; 3) the initial locations of the epidemics; and 4) the testing strategy that was implemented. We simulated epidemics for 72 scenarios by varying these four factors, and simulated 1,000 epidemics for each scenario over a 100-day period, as described in Section 1.1.3.

We chose a 100-day time period for the simulations under the assumption that effective vaccines or other pharmaceutical controls will be available after 100 days, in line with the missions of the Coalition for Epidemic Preparedness Innovations (CEPI) and other global health actors [4, 5]. The goal of decision-makers may therefore be to reduce the impacts of an epidemic prior to it reaching the 'controllable' stage at which vaccines or therapeutics become available. In the instance of KSA decision-makers, they may be interested in how they can minimise the health impacts of a potential epidemic without limiting pilgrim numbers. This simulation study showcased how the modelling framework can be used to compare the impacts and costs of different intervention scenarios.

| **Factor** | **Scenarios** |
|---|---|
| Movement input | MOH, IATA with baseline adjustment, IATA with no baseline adjustment |
| Pathogen | Influenza-X, SARS-CoV-X |
| Seed location | Global, Indonesia, United Kingdom |
| Testing strategy | No testing, 80% of pilgrims tested, 99% of pilgrims tested, Symptom-based testing strategy |

Table S1: **Factors influencing epidemics that were explored in the simulation study.** The scenarios that were considered under each factor are shown. See Sections 1.2.1 to 1.2.4 for more detail.

##### 1.2.1 Mobility proxies

In the absence of data on recent pilgrim movements, we considered three methods and data sources that could serve as proxies for estimating the numbers and origins of foreign pilgrims attending the Hajj. One approach was based on historical pilgrim data from a single Hajj published by Khan *et al* [6], and two approaches estimated pilgrim numbers using flight passenger data. The steps to generate each of these three estimates are outlined below. For all three methods, we generated the total pilgrim population by combining the estimates of foreign pilgrims with the number of domestic pilgrims that attended the Hajj in 2008 [7].

**Method 1: MOH data.** We used data from the KSA Ministry of Health (MOH) reported by Khan *et al* [6]. The dataset provided the pilgrim numbers from each of the 20 countries that sent the most foreign pilgrims to the Hajj in 2008, with the origin of an additional 301,634 foreign pilgrims listed as 'Other'. Hajj visas are in theory allocated to countries in proportion to the size of their Muslim population [8]. We therefore re-distributed the 'Other' category according to the estimated sizes of the Muslim populations in each of the countries not featured in the MOH top 20, using global Muslim population estimates from the Pew Center [9]. The approach we used to distribute the 'Other' category is similar to methods used in previous Hajj modelling work [10].

**Method 2: IATA with baseline adjustment.** We used flight passenger data purchased from IATA that contained the numbers of passengers that travelled between pairs of international airports each month in the period January 2012 to December 2021 [11]. To estimate the additional influx of passengers attributable to the Hajj, we adopted a simple approach to first estimate the baseline passenger level in the absence of the Hajj. We assumed that the minimum level of baseline travel corresponded to the month in the dataset in which there was no Hajj and the fewest passengers arrived in KSA (January 2012). An upper bound for the number of pilgrims could therefore be computed by subtracting the baseline from the passenger numbers in the month/year of the Hajj that had the highest total arrivals (August 2018). For each origin country, we estimated the number of pilgrims as *August2018_Arrivals – January2012_Arrivals*. Where this gave a negative number we set the pilgrim number equal to 0. To enable fair comparison between different pilgrim mobility proxies, we scaled the foreign pilgrim numbers estimated by this method so that the total number was equal to the total foreign pilgrims in the MOH dataset.

**Method 3: IATA with no baseline adjustment.** In this mobility proxy, we assumed that the origins of Hajj pilgrims were distributed in the same proportion as the origins of all flight passengers travelling to KSA. We used the number of flight passengers arriving in KSA in during the Hajj month/year used in Method 2 (August 2018). As in Method 2, we scaled the foreign pilgrim numbers estimated by this IATA method so that the total was equal to the total foreign pilgrims in the MOH dataset.

##### 1.2.2 Pathogen

We varied the natural history parameters in the model to simulate epidemics for two pathogen scenarios: i) a pathogen with similar characteristics to H1N1 Influenza that we label as 'Influenza-X'; ii) a pathogen with similar characteristics to SARS-CoV-2 that we label as a 'SARS-CoV-X'. The natural history parameters used for these two pathogen scenarios are shown in Table S2.

| **Parameter** | **Parameter value (Influenza-X scenario)** | **Parameter value (SARS-CoV-X scenario)** | **Source** |
|---|---|---|---|
| $\theta_a$ (relative infectiousness of asymptomatic)[1] | 0.58 | 0.58 | [12] |
| $\theta_p$ (relative infectiousness of presymptomatic)[1] | 1 | 1 | [13] |
| $\pi$ (proportion symptomatic) | 0.669 | 0.595 | [12] |

| | | | |
|---|---|---|---|
| $\delta$ (= 1 / mean duration $E$, $days^{-1}$) | 1/1.1 | 1/4.6 | [12] |
| $\gamma_p$ (= 1 / mean duration $I^p$, $days^{-1}$) | 1/1.0 | 1/1.7 | [12, 13] |
| $\gamma_s$ (= 1 / mean duration $I^s$, $days^{-1}$) | 1/1.5 | 1/2.1 | [12, 13] |
| $\gamma_a$ (= 1 / mean duration $I^a$, $days^{-1}$) | 1/2.5 | 1/2.1 | [12] |
| $R_0$ (basic reproduction number) | 1.64 | 2.66 | [12] |

Table S2: **Parameter values for the Influenza-X and SARS-CoV-X scenarios.** We selected values for $\beta$ to give $R_0$ values that were in line with a range of previously estimated reproduction numbers for Influenza and SARS-CoV epidemics. [[1] Relative to infectiousness of symptomatic individuals.]

##### 1.2.3 Seed location

We simulated three outbreak location scenarios: i) an emerging epidemic that was localised in Indonesia (the largest source of foreign pilgrims) at the time of the Hajj; ii) an emerging epidemic that was localised in the United Kingdom (selected as an example country that is a small source of foreign pilgrims) at the time of the Hajj; iii) epidemics were established in multiple countries and continents at the time of the Hajj (this scenario is referred to as 'Global' throughout).

In scenarios i) and ii), we began the simulations with 1,000 individuals infected in Indonesia and the United Kingdom respectively. This meant that the initial infection prevalence was much lower in the Indonesian scenario than the United Kingdom scenario, but may correspond to similarly early stages of an outbreak when it is localised within a particular town or city rather than countrywide (such as in the early stages of the COVID pandemic when the outbreak was centred in Wuhan).

In the Global scenario, we seeded infections in the three most populated countries in each of the six World Health Organization regions (Section 1.5). We began the simulations with 0.001% of individuals infected in each of the 18 selected countries. In comparison to the local, emerging epidemics of scenarios i) and ii), the Global scenario allowed me to explore a situation with an established pandemic.

##### 1.2.4 Testing strategy

We explored four different testing strategies in the simulations: i) No testing; ii) Randomly testing 80% of arriving pilgrims; iii) Randomly testing 99% of arriving pilgrims; iv) a symptom-based testing strategy (also referred to as 'syndromic').

In the 80% and 99% testing strategies, we set $\tau = 0.80$ or 0.99 in Eq. (S7). We assumed a rapid antigen test was used with sensitivity of 0.7680 and a specificity of 0.9968 based on a SARS-CoV-2 test [14].

In the syndromic testing strategy, we assumed that there was a preliminary temperature scanning step, as has been introduced in response to some previous epidemics [15]. Any pilgrims identified as having an elevated temperature would undergo a further test. We assumed that this step would identify 87% of infected pilgrims and 39% of uninfected pilgrims for further testing, based on previous estimates of the effectiveness of airport thermal screening for influenza [16]. The pilgrims identified for further testing by thermal screening were then all tested with a rapid antigen test (same sensitivity and specificity as detailed above in the 80 and 99% testing strategies [14]). The steps for syndromic testing can be summarised as:

$$\begin{aligned}
&X^{tested}_{i\to KSA} \sim Binomial\left(X^{travel}_{i\to KSA}, \tau^{X}_{therm}\right)\\
&\text{where } X = S, E, I^a, I^p, I^s, R\\
\\
&\text{When } X = E, I_a, I_p \text{ or } I_s\text{:}\\
&X^{diagnosed}_{i\to KSA} \sim Binomial\left(X^{tested}_{i\to KSA}, \epsilon\right)\\
\\
&\text{When } X = S \text{ or } R\text{:}\\
&X^{false_diagnosed}_{i\to KSA}(t) \sim Binomial\left(X^{tested}_{i\to KSA}, \mu\right)
\end{aligned} \tag{S12}$$

where $\tau^{X}_{therm}$ is the detection rate for thermal screening, taking the value 0.87 when $X = E, I_a, I_p$ or $I_s$, and 0.39 when $X = S$ or $R$. $\epsilon$ is the test sensitivity of the follow-up test, and $\mu$ represents the false positive rate, calculated as 1 - test specificity.

##### 1.2.5 Summary metrics for simulations

We assessed the outcomes of each simulated epidemic using the following model outputs:

- Epidemiological outcomes, including:
  - Total cases in pilgrims (foreign and domestic) and KSA non-pilgrims
  - Hospitalisations and deaths: To compute these, we first distributed cases into age and sex categories using estimates of age and sex distributions for the different populations in the model. Detailed demographic data by pilgrim nationality were unavailable, so we assumed that the ages and sex of all foreign pilgrims followed distributions reported by Azarpazhooh *et al* for a large cohort of 92,974 Iranian pilgrims (with age categories 15–24, 25–34, 35–44, 45–54, 55–64, 65–74, 75–84, $> 85$) [17]. For domestic non-pilgrims, we used United Nations World Population Prospects (UN WPP) data for KSA with the same age categories as above, plus a category for $< 15$ [18]. For domestic pilgrims, we modified the UN WPP age distribution to exclude the $< 15$ category. This assumption that children

would not attend the Hajj has been used in previous Hajj models [19]. We then applied age-specific infection hospitalisation ratios (IHR) and infection fatality ratios (IFR) to convert the cases by age group into estimates of hospitalisations and deaths. We used different IHR and IFR distributions for the Influenza X and SARS-CoV-X scenarios (Table S4).

  - Years of life lost (YLL): we assumed that ages were distributed uniformly within age categories and that the mean age of death equalled the mean age of the age category. For each value of age ($a$) and sex ($s$) we computed:

$$YLL_{a,s} = D_{a,s}L_{a,s} \tag{S13}$$

    where $D_{a,s}$ is the number of deaths for the given age $a$ and sex $s$, and $L_{a,s}$ is the remaining life expectancy at the age at which death occurs. It is typical to use life expectancies at each age based on an ideal standard, so we used life expectancies from Japan, a country with one of the highest life expectancies [20–22].

- Economic outcomes, including:
  - Scenario costs: We searched the literature for cost estimates of the components of the intervention strategies. Data from KSA were scarce for many of the intervention components so we generated estimates of up-to-date costs in KSA based on estimates from other countries. We first converted foreign costs to Saudi Riyals using historical exchange rates [23]. We then obtained a 2023 cost estimate by adjusting for inflation using historical consumer price index data from KSA before converting costs into international dollars [24, 25]. The component cost estimates are summarised in Table S3 and were used to estimate the total costs of each scenario (including the costs of the testing interventions and hospital treatment).

| **Item** | **Cost** | **Source** | **Converted and inflation-adjusted costs (2023 international $)** |
|---|---|---|---|
| Cost of running an airport screening programme | AU$0.24 / passenger (2009) (based on reported costs of AU$150,000 for 625,147 passengers screened) | [26] | Intl$0.52 / passenger screened |
| Cost per diagnostic test | US$5 (2020) | [27] | Intl$10.61 |
| Cost of quarantine per patient | US$1,071 (2020) | [28] | Intl$2,272.69 |
| Treatment cost per hospitalised patient in KSA | US$10,182.50 (2020) (based on estimates of average SARS-CoV-2 treatment costs in KSA) | [29] | Intl$21,607.48 |

Table S3: Cost estimates for various health interventions with inflation-adjusted values in 2023 international dollars

– Net monetary benefits of testing strategies. The net monetary benefit (NMB) compares the costs and benefits of an intervention assuming a willingness to pay (WTP) threshold for a unit of health benefit [30]. The NMB was calculated as:

$$NMB = (T\Delta B) - \Delta C \tag{S14}$$

where $T$ is the willingness to pay threshold, $\Delta B$ is the YLL averted by the intervention, and $\Delta C$ is the cost of the intervention scenario minus the cost of the counterfactual scenario of no testing. We used WTP thresholds per Life Year estimated for KSA by Pichon-Riviere *et al* which we converted to 2023 international dollars [31]. We estimated NMBs using the central estimate of the KSA WTP threshold (Intl$20,324 for a YLL averted), and performed sensitivity analyses using the lower and upper limits of their estimated WTP range (Intl$17,298 and Intl$25,452 respectively). We also evaluated how the probability that an intervention was deemed to be cost-effective (NMB ¿ 0) varied against the WTP threshold.

For each scenario, we summarised the distributions of each outcome across all 1,000 simulations using the median, 2.5%, 25%, 75%, and 97.5% quantiles. We paired simulations between the intervention (i.e. with testing) and counterfactual (i.e. no testing) scenarios to compute differences in measures such as YLL and costs. The results presented in the main text are based on a ranked pairing method, whereby for each intervention scenario we first ordered the 1,000 simulations by the total number of cases occurring in KSA. We then paired each of the ranked simulations with the equal ranking simulation from the corresponding baseline scenario (matched on movement input, pathogen, and seed location). This approach was taken in order to compare epidemic trajectories to suitable counterfactuals, and limit misleading conclusions on the effects of interventions that can arise from pairing based on matching by initial random seeds [32]. In the Supplementary Results below we include results from using a pairing method where the simulations were matched based on the value of an initial random seed within the simulation code.

#### 1.3 Software

All epidemic simulations and analyses were performed using the R software [33]. The model code is available in the *multipatchr* package in R (`https://github.com/sangeetabhatia03/multipatchr`). The code for the analysis in this paper is available at `https://github.com/j-wardle/hajj_modelling_study`.

#### 1.4 Hospitalisation and fatality ratios for pathogen scenarios

| | SARS-CoV-X | | Influenza X | |
|---|---|---|---|---|
| Age group (years) | IHR | IFR | IHR | IFR |
| 0-4 | 0.0000160 | 0.0000160 | 0.00466 | 0.000185 |
| 5-9 | 0.0000160 | 0.0000160 | 0.00183 | 0.0000736 |
| 10-14 | 0.000408 | 0.0000700 | 0.00183 | 0.0000736 |
| 15-19 | 0.000408 | 0.0000700 | 0.00183 | 0.0000803 |
| 20-24 | 0.0104 | 0.000309 | 0.00183 | 0.0000803 |
| 25-29 | 0.0104 | 0.000309 | 0.00375 | 0.000201 |
| 30-34 | 0.0343 | 0.000844 | 0.00375 | 0.000201 |
| 35-39 | 0.0343 | 0.000844 | 0.00375 | 0.000201 |
| 40-44 | 0.0425 | 0.00161 | 0.00375 | 0.000201 |
| 45-49 | 0.0425 | 0.00161 | 0.00375 | 0.000435 |
| 50-54 | 0.0816 | 0.00595 | 0.00709 | 0.000435 |
| 55-59 | 0.0816 | 0.00595 | 0.00709 | 0.000435 |
| 60-64 | 0.118 | 0.0193 | 0.00709 | 0.000435 |
| 65-69 | 0.118 | 0.0193 | 0.0103 | 0.00656 |
| 70-74 | 0.166 | 0.0428 | 0.0103 | 0.00656 |
| 75-79 | 0.166 | 0.0428 | 0.0103 | 0.00656 |
| >80 | 0.184 | 0.0780 | 0.0103 | 0.00656 |

Table S4: **Infection hospitalisation ratios (IHR) and infection fatality ratios (IFR) by age group for computing hospitalisations and deaths in the SARS-CoV-X and Influenza X simulation scenarios.** Values are provided to 3 significant figures and are based on a review of pandemic pathogen characteristics in work by Doohan *et al* [12].

#### 1.5 Seeding scenarios

In the Global scenario we seeded infections in the three most populated countries in each of the six World Health Organisation regions. These were:

- **African region:** Nigeria, Ethiopia, Democratic Republic of the Congo;
- **Region of the Americas:** United States of America, Brazil, Mexico;
- **Eastern Mediterranean Region:** Egypt, Pakistan, Iran;
- **European Region:** Russian Federation, Germany, Turkey;
- **South-East Asia Region:** India, Indonesia, Bangladesh;
- **Western Pacific Region:** China, Philippines, Vietnam.

### 2 Supplementary results

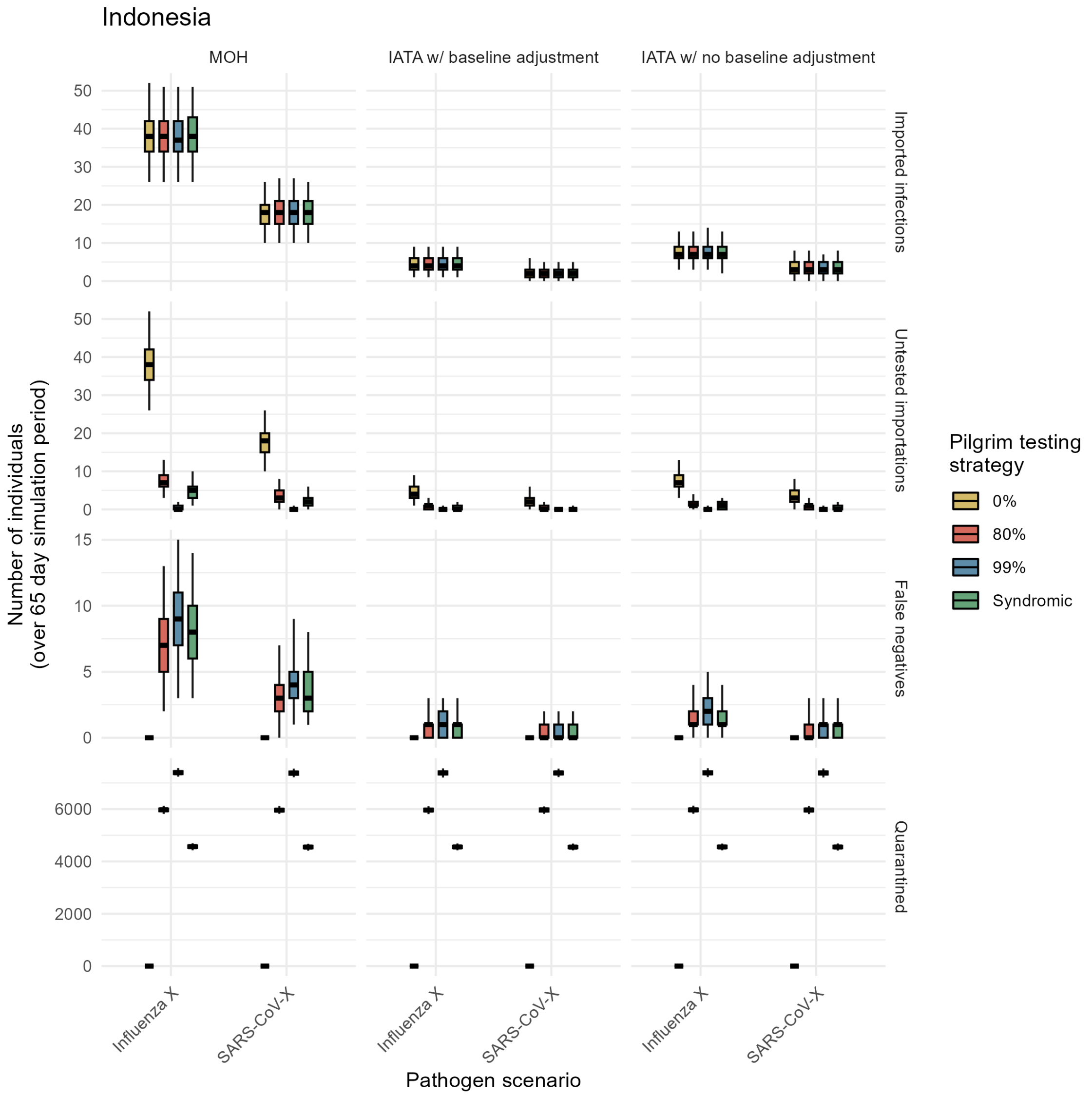

Figure S2: **Testing outcomes by movement input method for simulations in the Indonesia seeding scenario.** Values are presented by movement input method (columns) and pathogen (x-axis). Colours denote the testing strategy. Box and whisker plots summarise the median (thick black horizontal line), interquartile range (coloured rectangle) and 95% quantiles (range denoted by black lines) across 1,000 simulations.

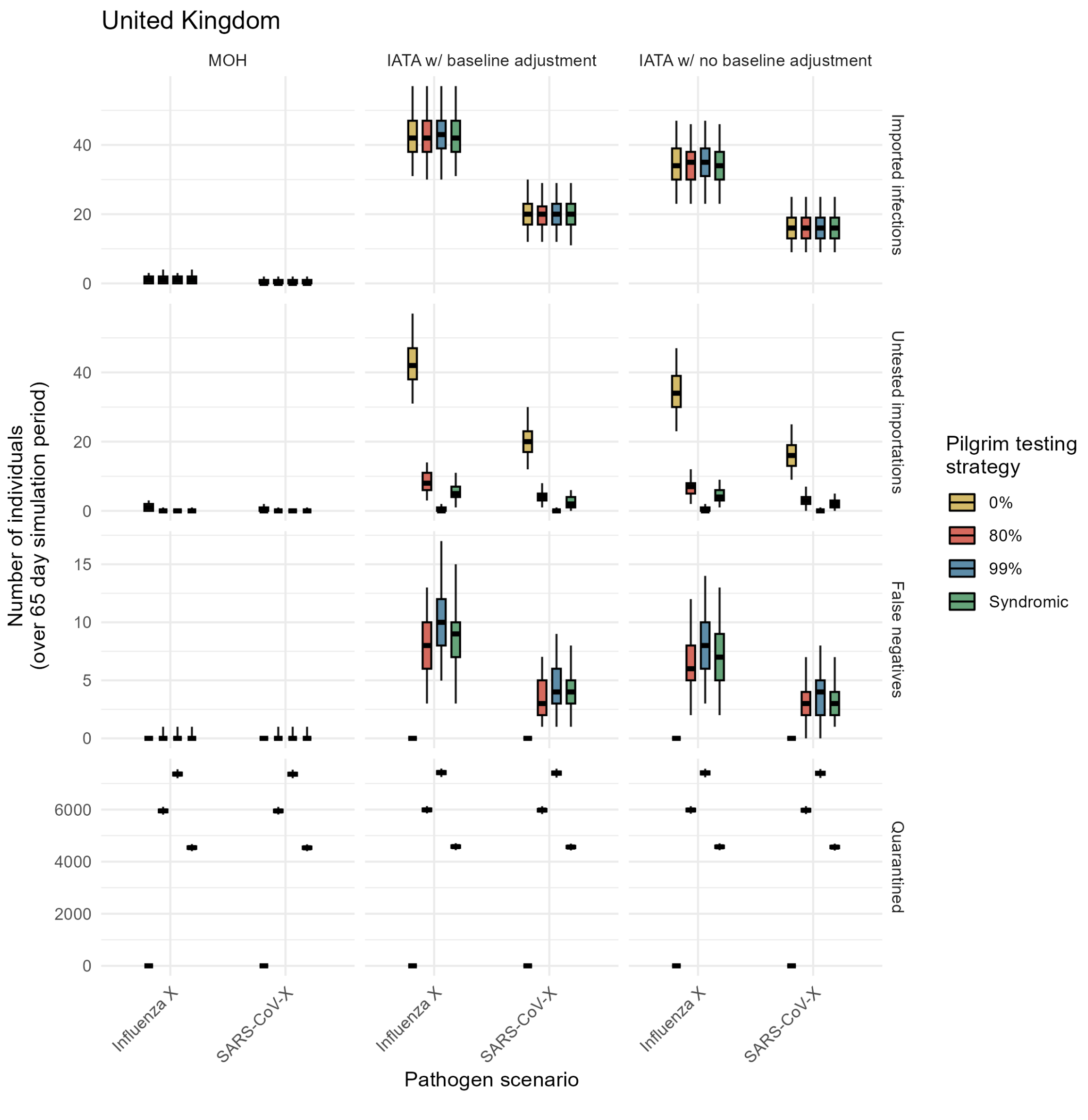

Figure S3: **Testing outcomes by movement input method for simulations in the United Kingdom seeding scenario.** Caption as Fig. S2.

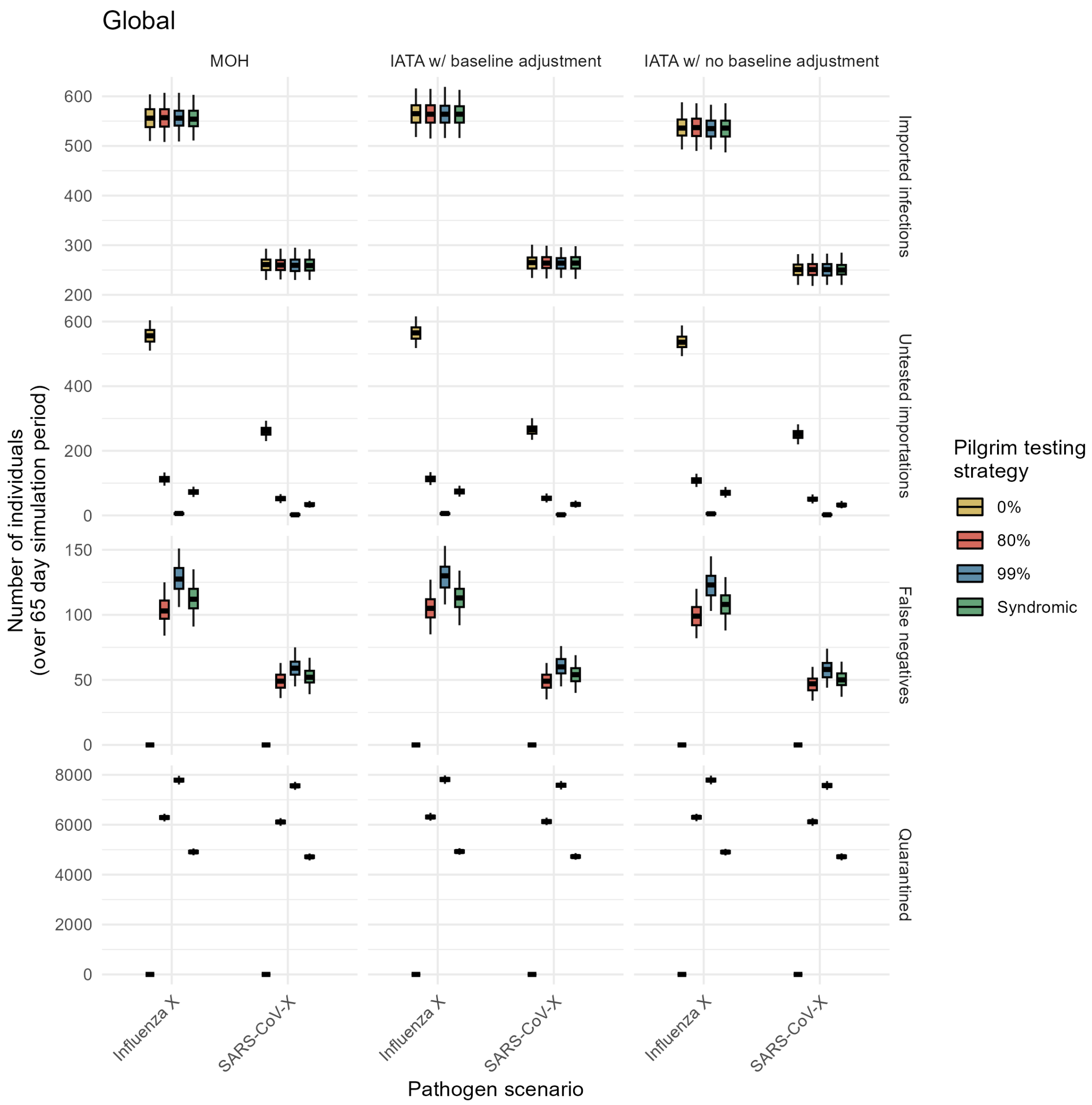

Figure S4: **Testing outcomes by movement input method for simulations in the Global seeding scenario.** Caption as Fig. S2.

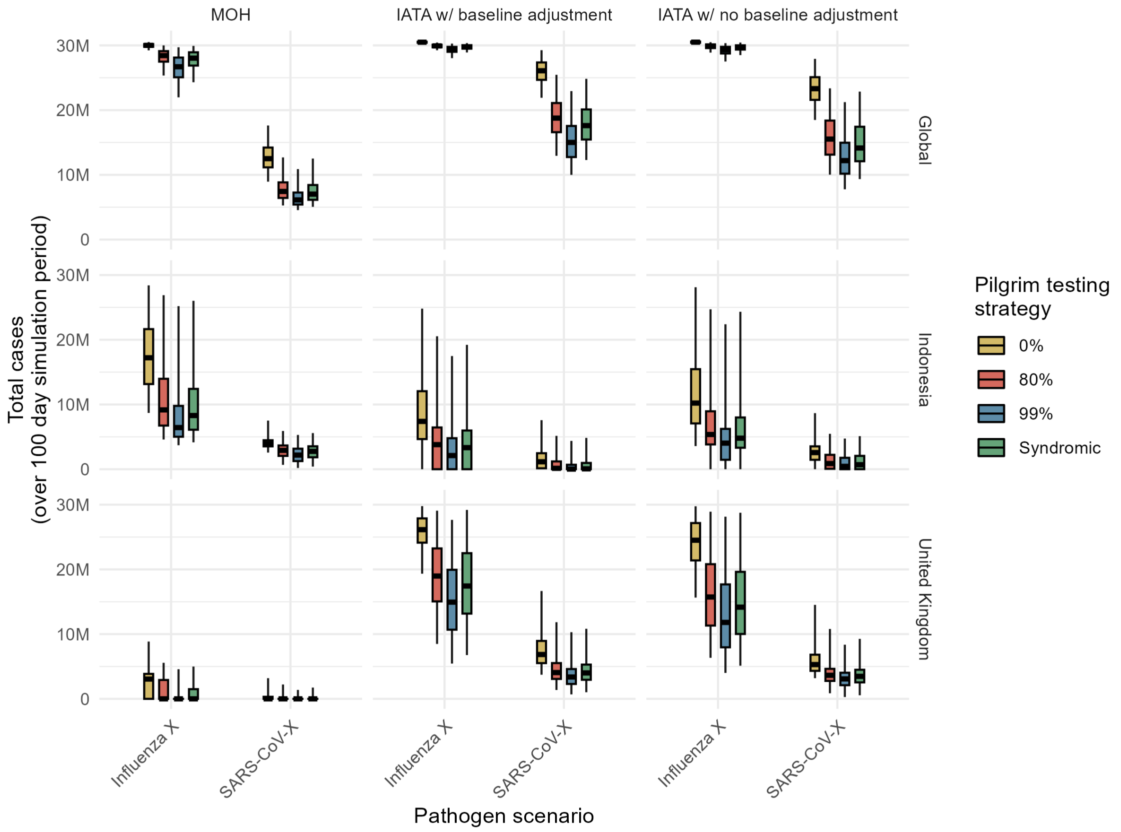

Figure S5: **Total cases in KSA by movement input method.** Values for the total cases in KSA (comprised of KSA residents and pilgrims infected while in KSA) are presented by movement input method (columns), epidemic seed scenarios (rows), and pathogen (x-axis). Colours denote the testing strategy. Box and whisker plots summarise the median (thick black horizontal line), interquartile range (coloured rectangle) and 95% quantiles (range denoted by black lines) across 1,000 simulations.

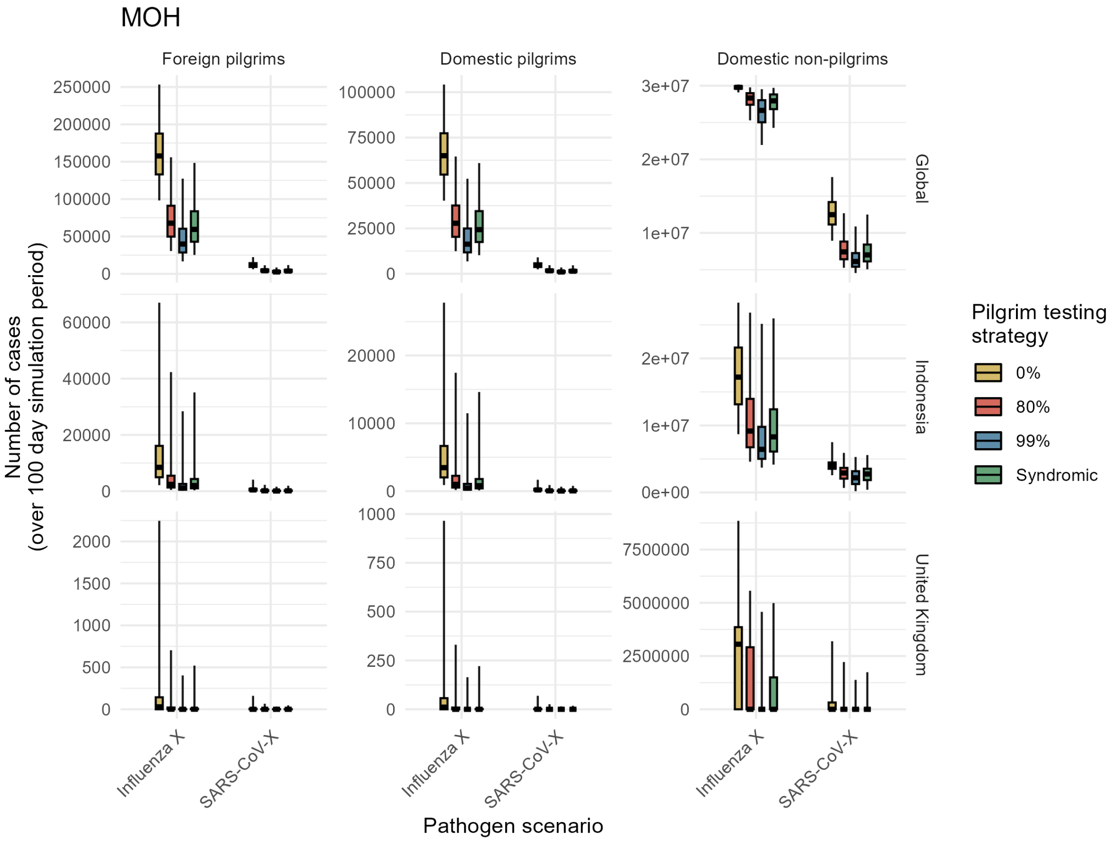

Figure S6: **Total cases by pilgrim sub-population for simulations using MOH movement inputs.** Values are presented by pilgrim sub-population (columns), seeding location (rows), and pathogen (x-axis). Colours denote the testing strategy. Box and whisker plots summarise the median (thick black horizontal line), interquartile range (coloured rectangle) and 95% quantiles (range denoted by black lines) across 1,000 simulations.

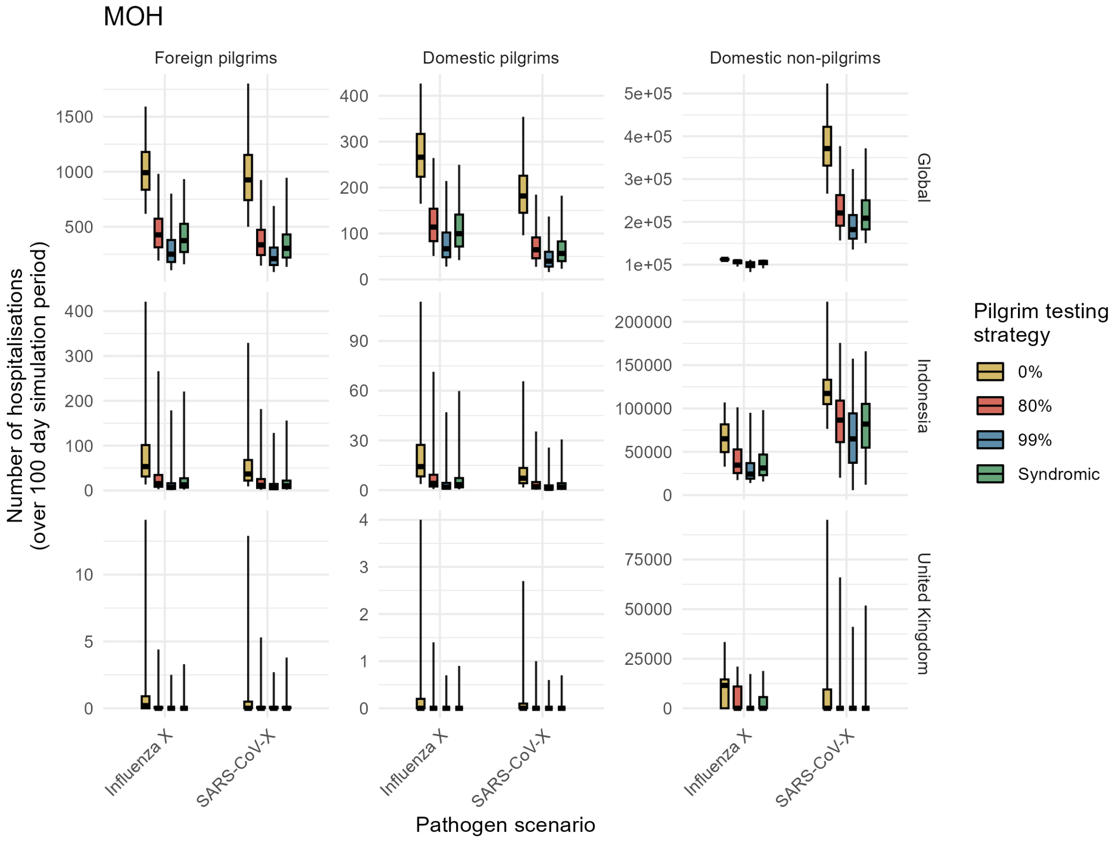

Figure S7: **Total hospitalisations by pilgrim sub-population for simulations using MOH movement inputs.** Caption as Fig. S6.

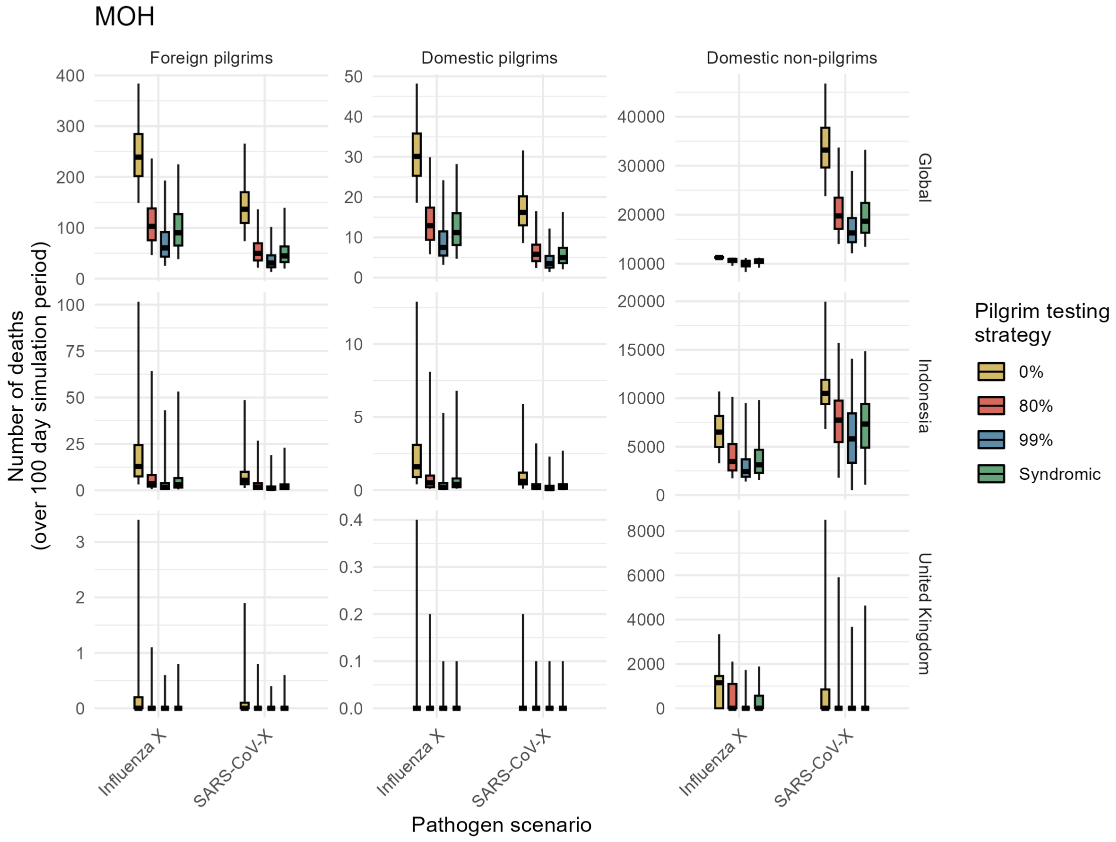

Figure S8: **Total deaths by pilgrim sub-population for simulations using MOH movement inputs.** Caption as Fig. S6.

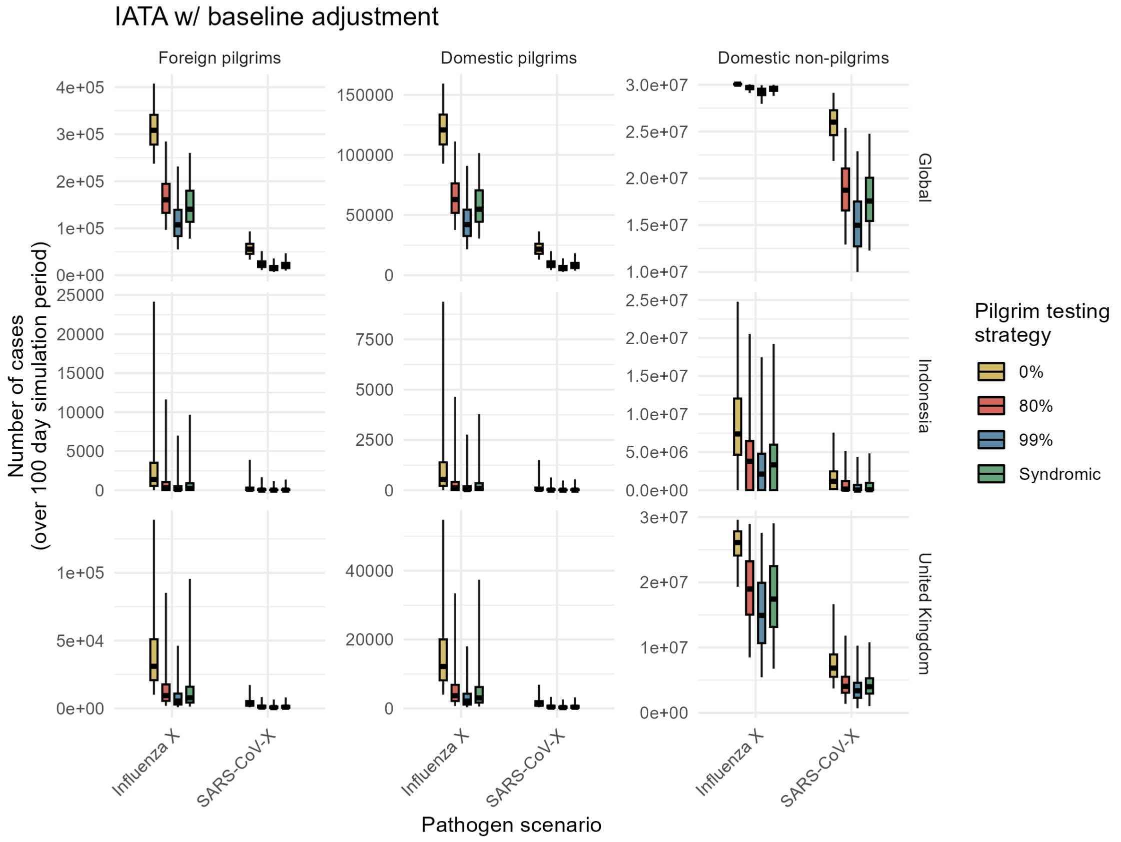

Figure S9: **Total cases by pilgrim sub-population for simulations using baseline-adjusted IATA movement inputs.** Caption as Fig. S6.

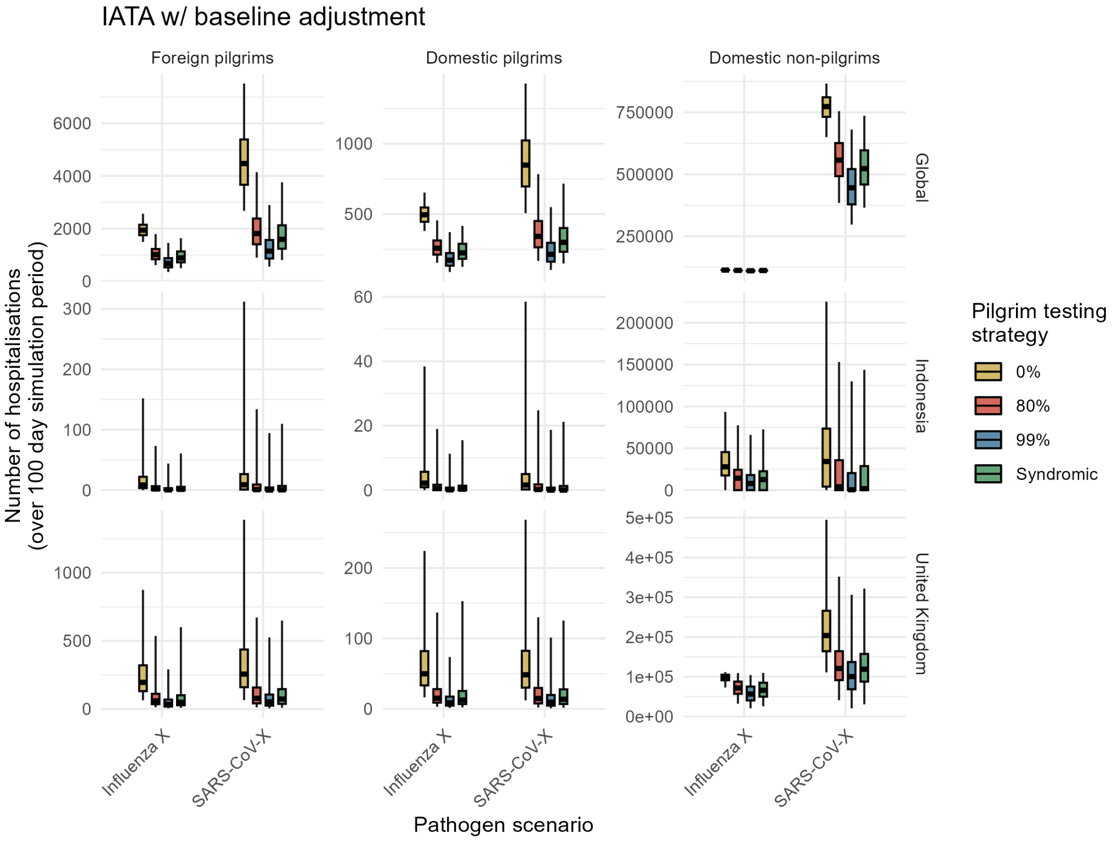

Figure S10: **Total hospitalisations by pilgrim sub-population for simulations using baseline-adjusted IATA movement inputs.** Caption as Fig. S6.

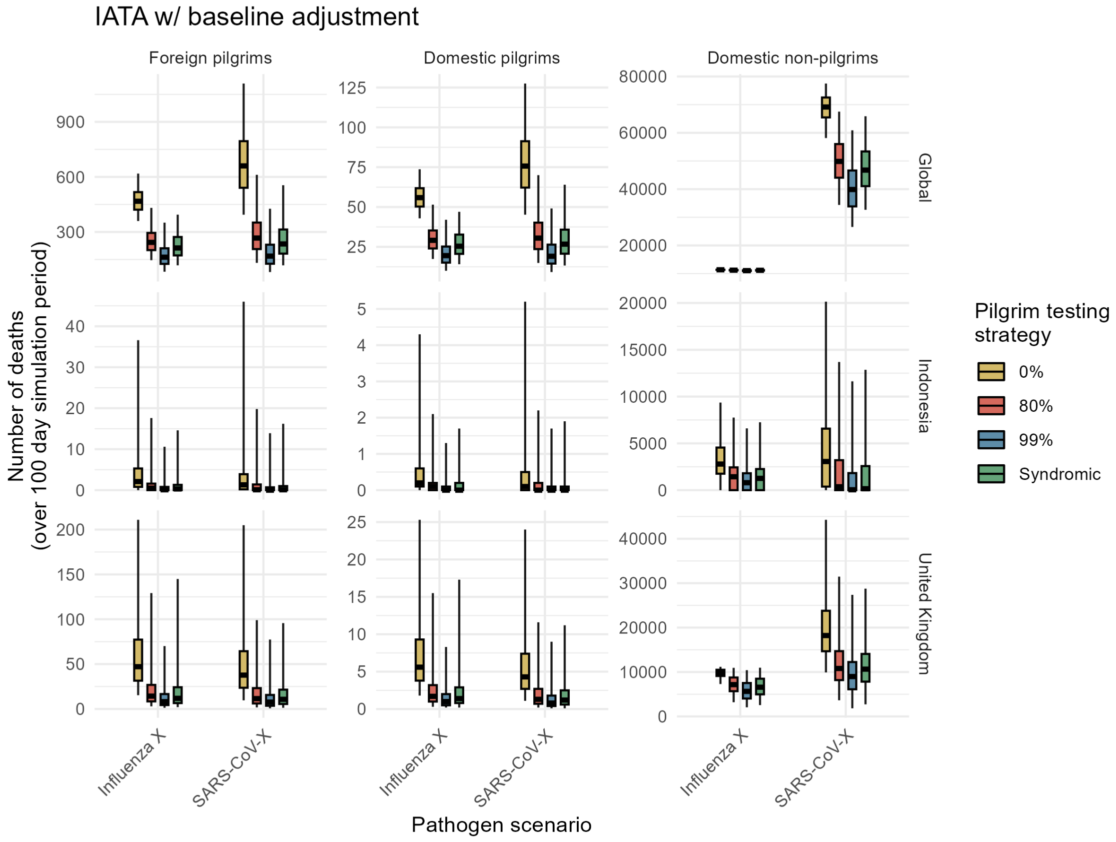

Figure S11: **Total deaths by pilgrim sub-population for simulations using baseline-adjusted IATA movement inputs.** Caption as Fig. S6.

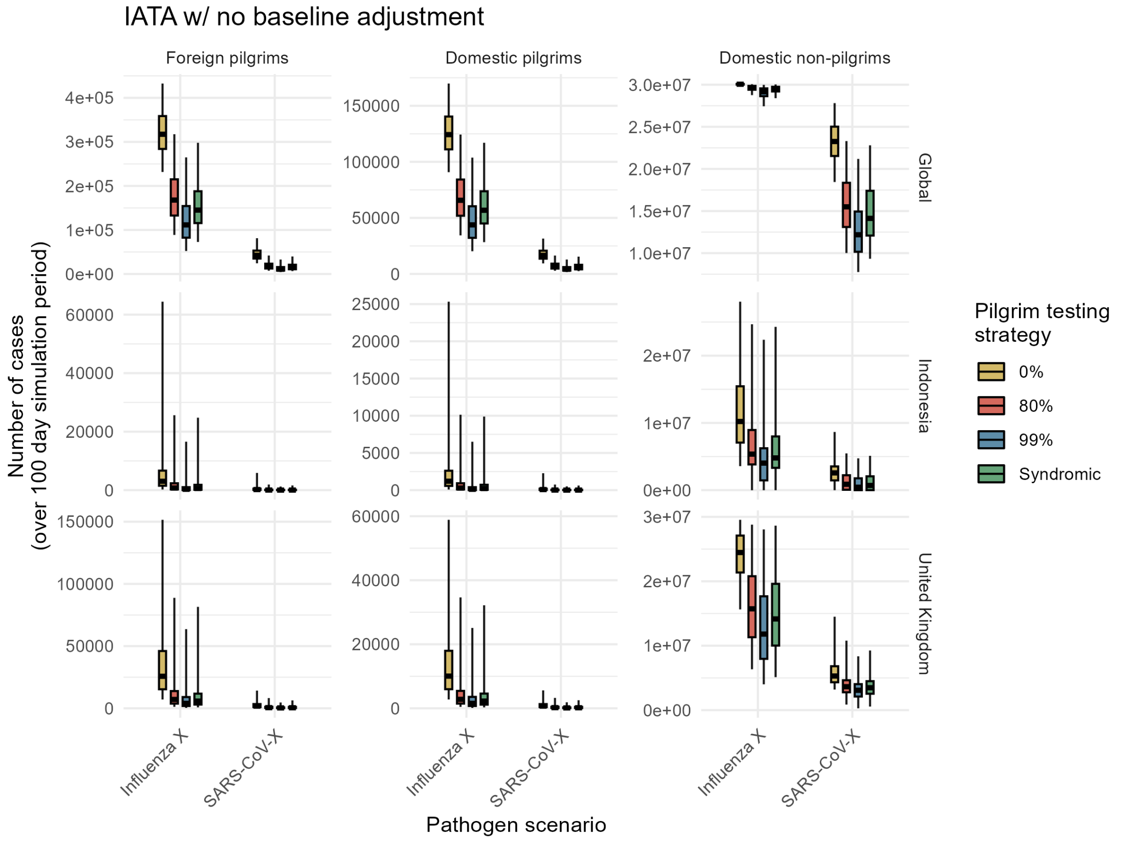

Figure S12: **Total cases by pilgrim sub-population for simulations using non baseline-adjusted IATA movement inputs.** Caption as Fig. S6.

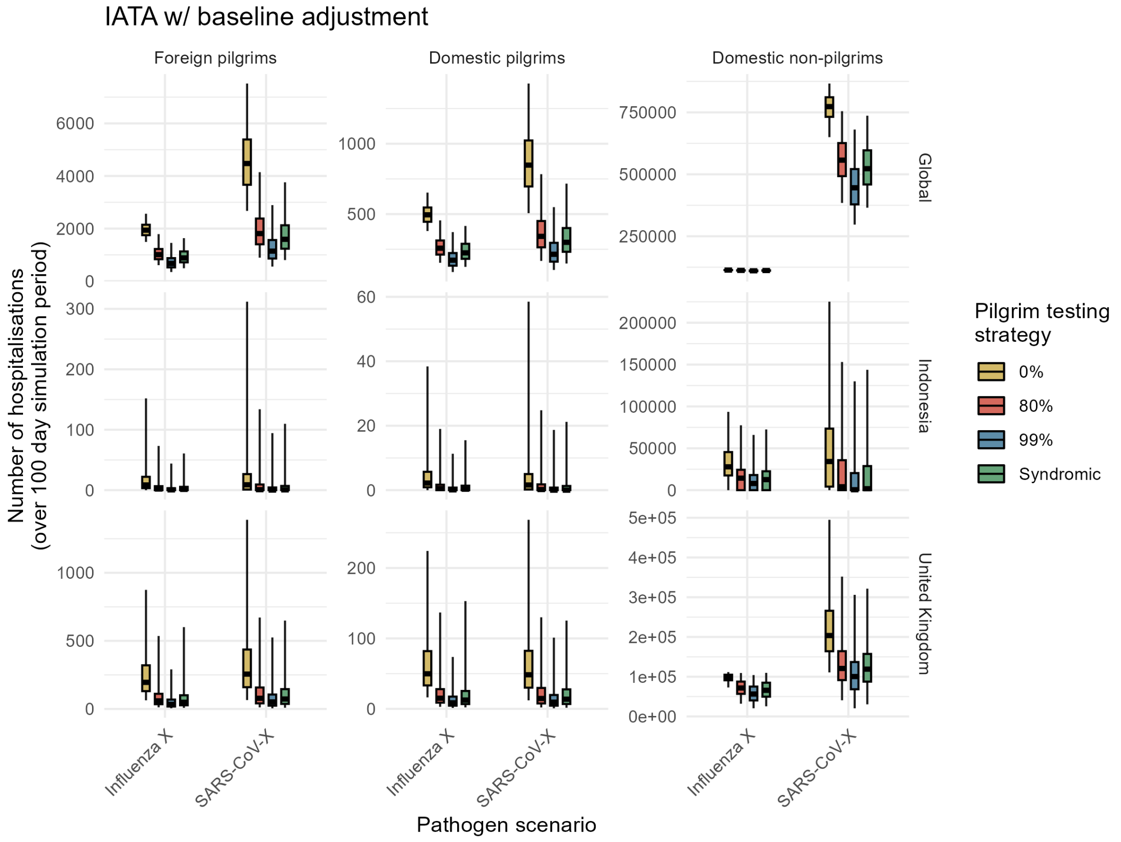

Figure S13: **Total hospitalisations by pilgrim sub-population for simulations using non baseline-adjusted IATA movement inputs.** Caption as Fig. S6.

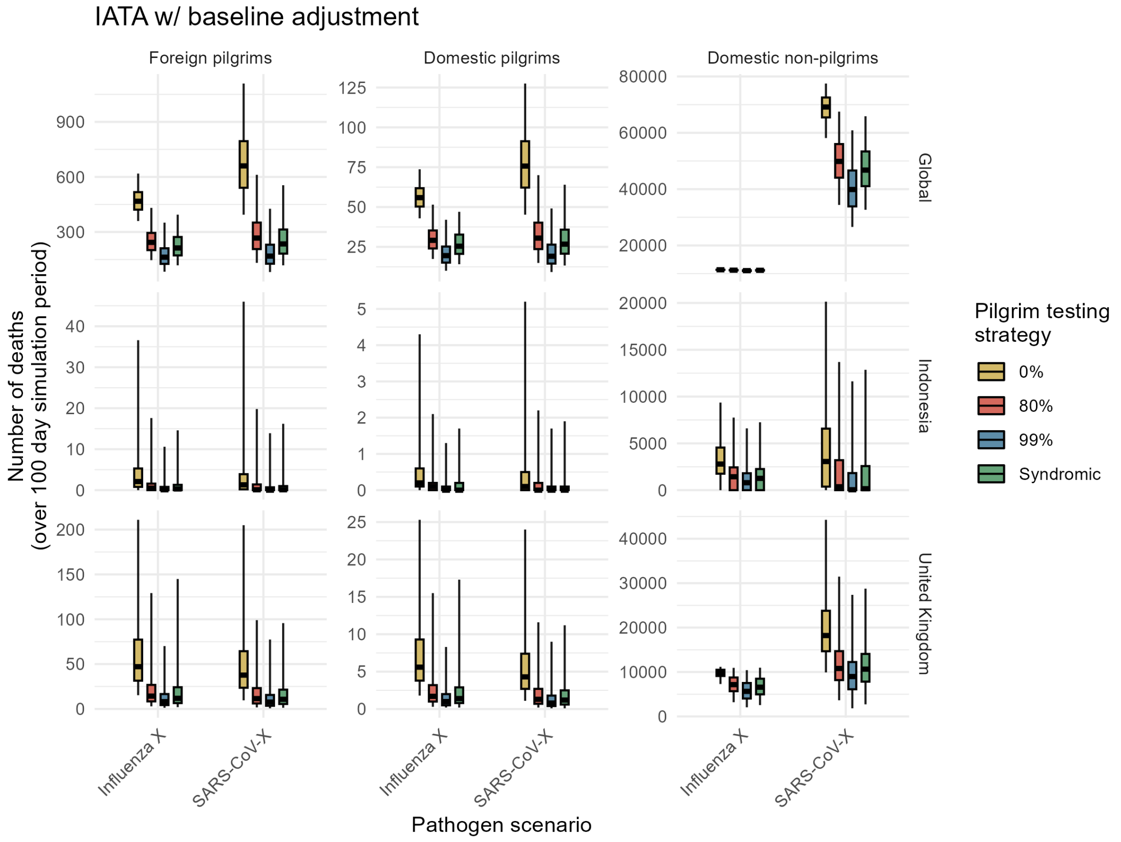

Figure S14: **Total deaths by pilgrim sub-population for simulations using non baseline-adjusted IATA movement inputs.** Caption as Fig. S6.

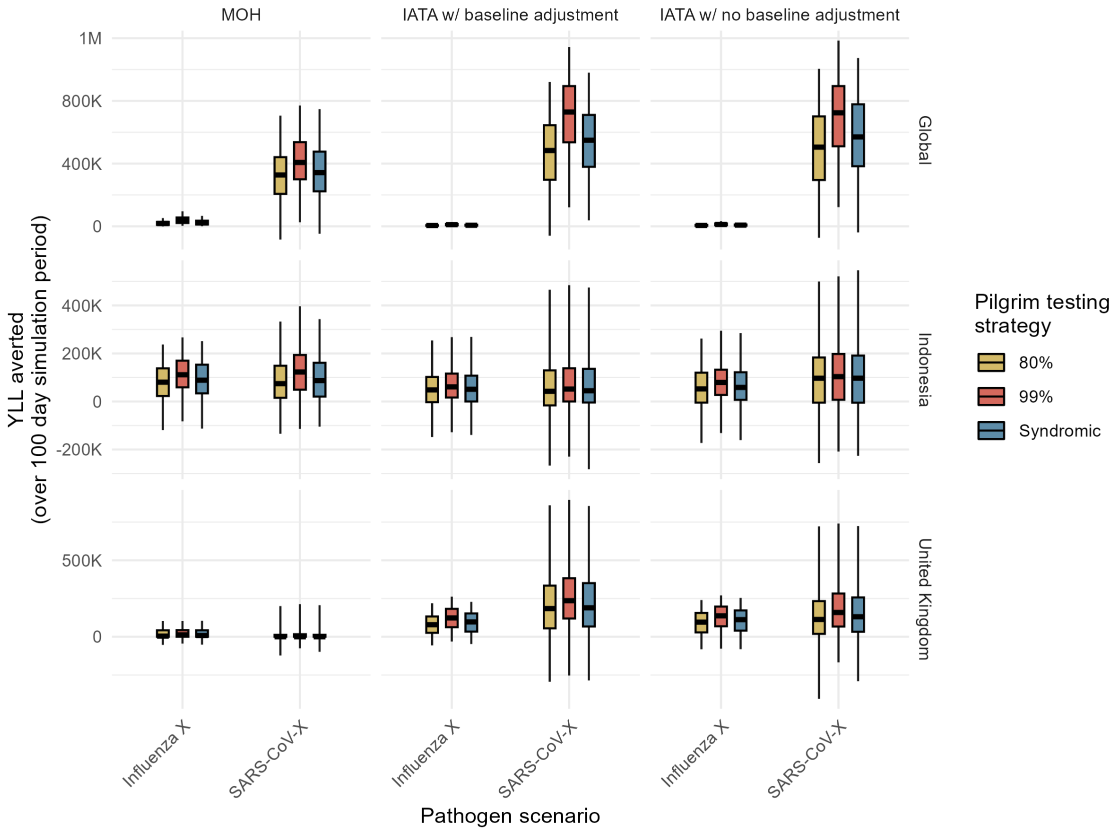

Figure S15: **Years of life lost averted through testing interventions, by movement input method (for simulations paired using the matched seed method).** Values are presented by pilgrim sub-population (columns), seeding location (rows), and pathogen (x-axis). Colours denote the testing strategy. Box and whisker plots summarise the median (thick black horizontal line), interquartile range (coloured rectangle) and 95% quantiles (range denoted by black lines) across 1,000 simulations.

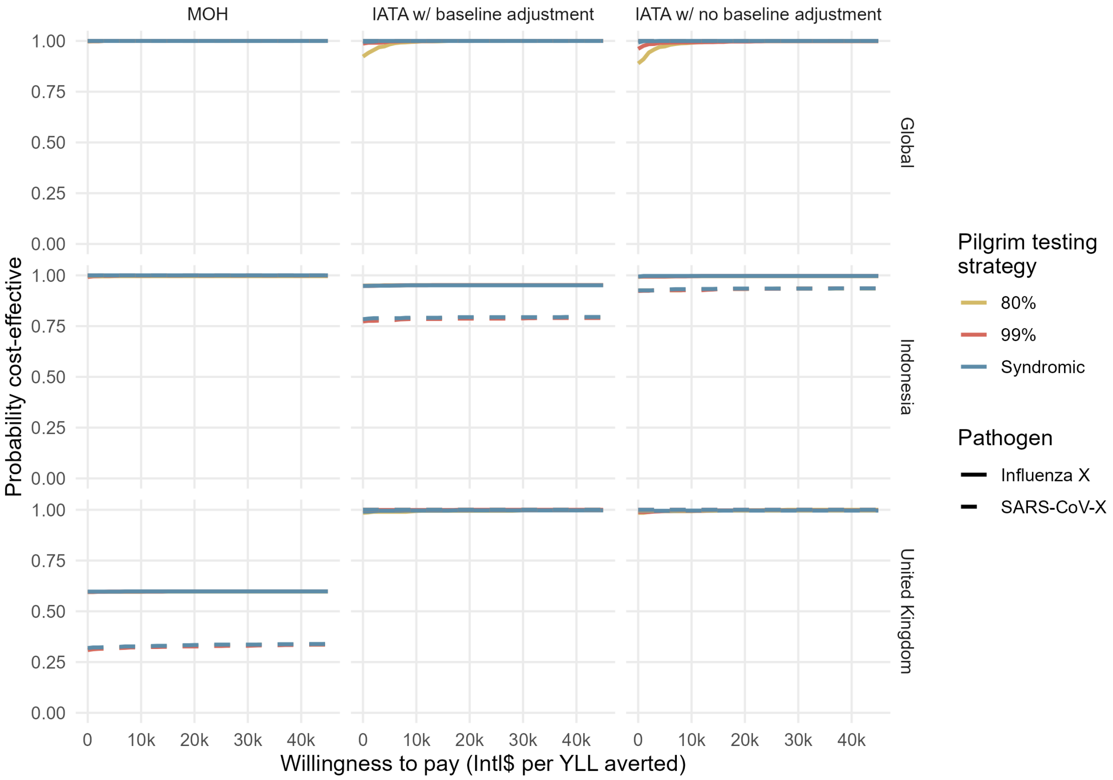

Figure S16: **Probability that interventions are cost-effective vs willingness to pay values.** Probabilities are based on the proportion of 1,000 sim pairings where the intervention strategy was evaluated to be cost-effective (net monetary benefits > Intl$0). Values are presented by seeding location (rows) and method used to estimate pilgrim numbers (columns). Colours denote the different strategies. Dashed lines = SARS-CoV-X, solid lines = Influenza X.

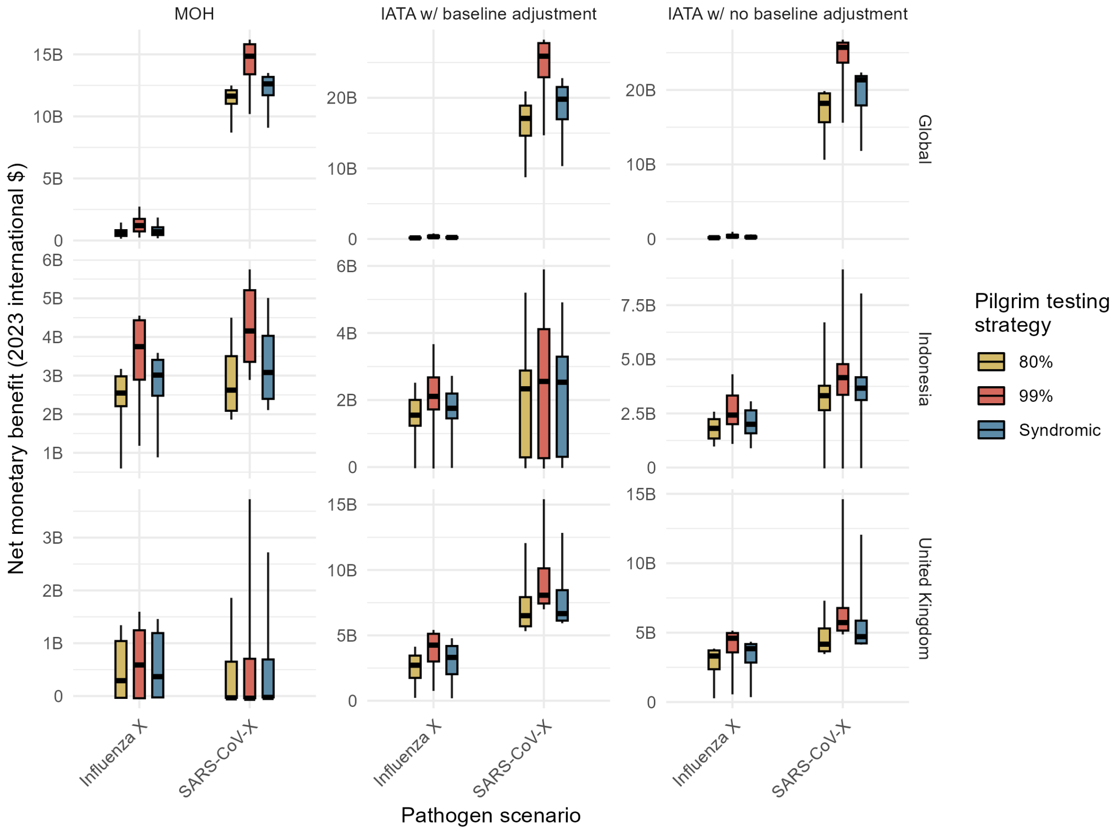

Figure S17: **Net monetary benefit estimates for different intervention scenarios, relative to a baseline scenario with no testing - sensitivity analysis using a higher willingness to pay threshold (Intl$25,452 for a YLL averted, based on values reported by Pichon-Riviere *et al* [31]).** Caption as Fig. S6.

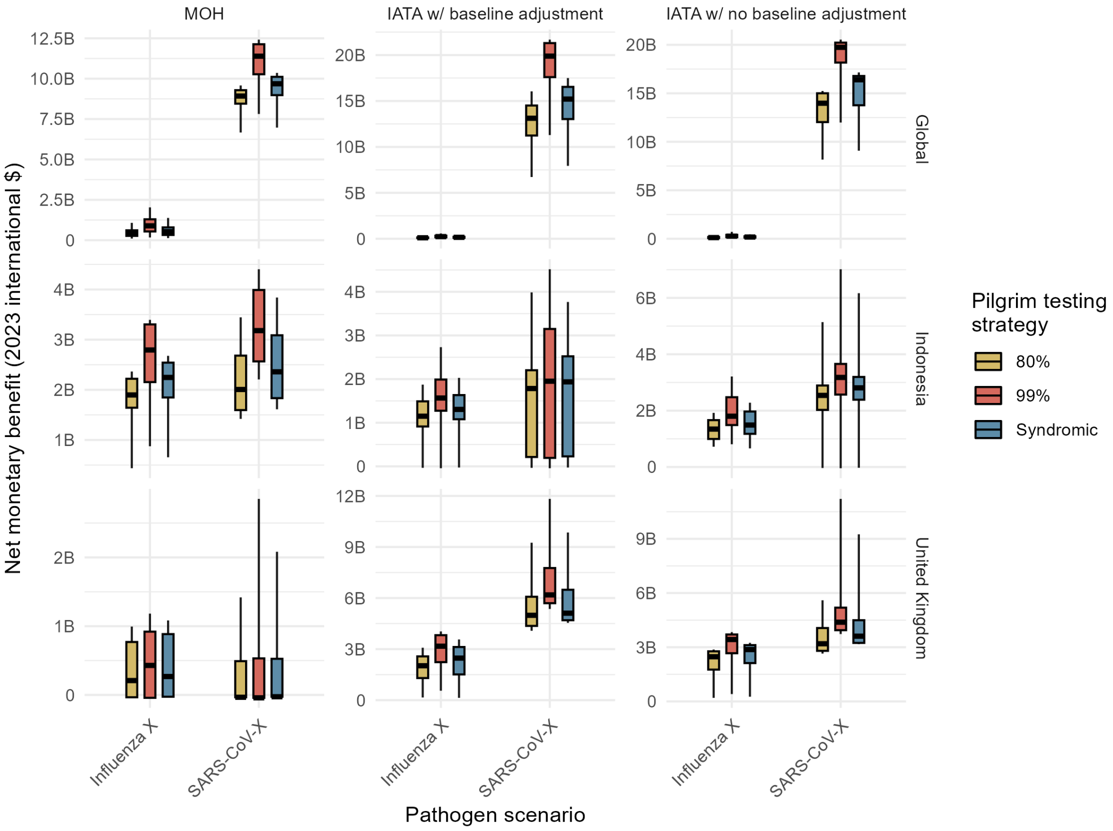

Figure S18: **Net monetary benefit estimates for different intervention scenarios, relative to a baseline scenario with no testing - sensitivity analysis using a lower willingness to pay threshold (Intl$17,298 for a YLL averted), based on values reported by Pichon-Riviere *et al* [31]).** Caption as Fig. S6.

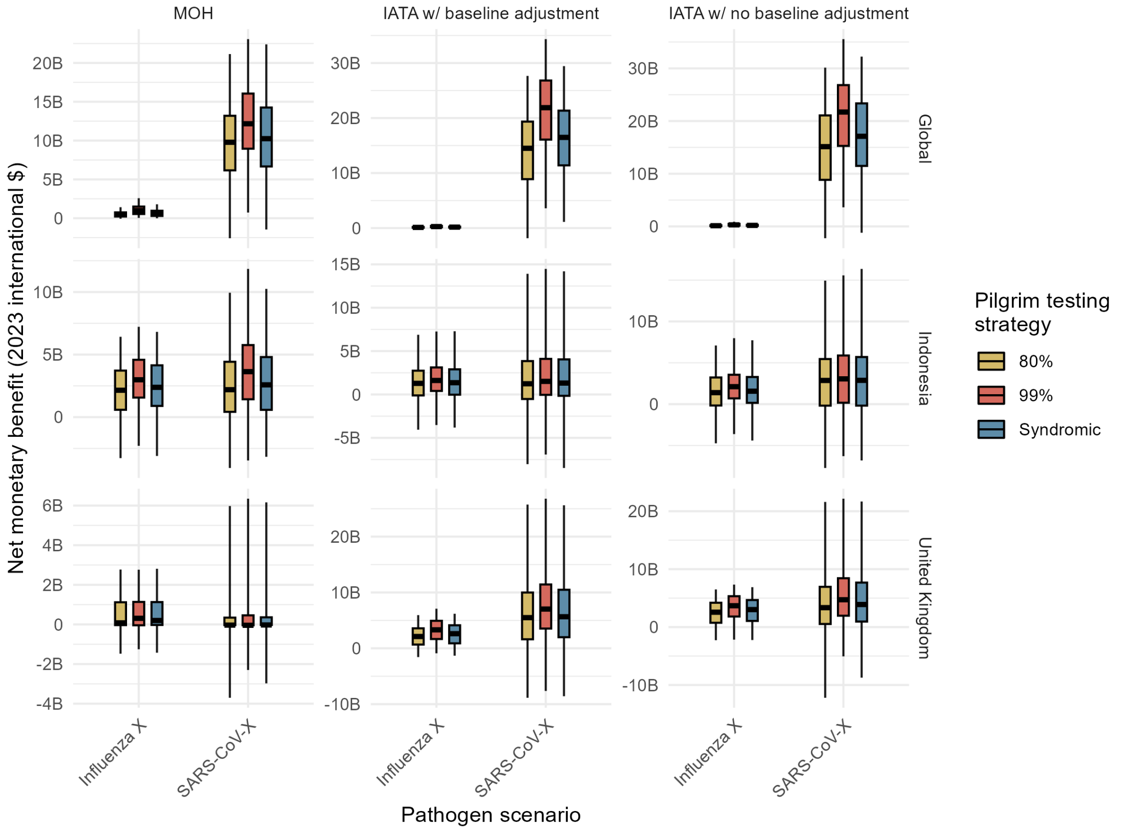

Figure S19: **Net monetary benefit estimates for different intervention scenarios, relative to a baseline scenario with no testing (for simulations paired using the matched seed method).** Caption as Fig. S6.

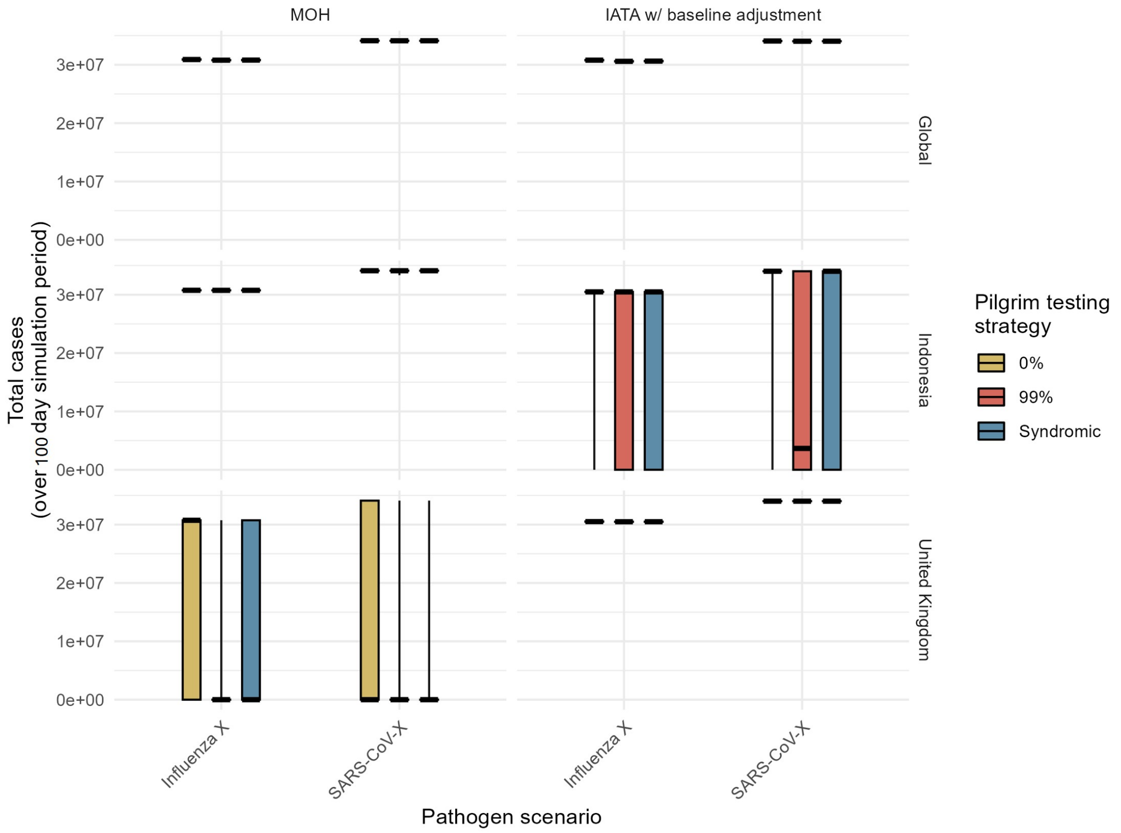

Figure S20: **Total cases in KSA under a 300-day simulation period.** Values for the total cases in KSA (comprised of KSA residents and pilgrims infected while in KSA) are presented by movement input method (columns), epidemic seed scenarios (rows), and pathogen (x-axis). Colours denote the testing strategy. Box and whisker plots summarise the median (thick black horizontal line), interquartile range (coloured rectangle) and 95% quantiles (range denoted by black lines) across 1,000 simulations.
